# Improving Polygenic Prediction in Ancestrally Diverse Populations

**DOI:** 10.1101/2020.12.27.20248738

**Authors:** Yunfeng Ruan, Yen-Feng Lin, Yen-Chen Anne Feng, Chia-Yen Chen, Max Lam, Zhenglin Guo, Stanley Global Asia Initiatives, Lin He, Akira Sawa, Alicia R. Martin, Shengying Qin, Hailiang Huang, Tian Ge

## Abstract

Polygenic risk scores (PRS) have attenuated cross-population predictive performance. As existing genome-wide association studies (GWAS) were predominantly conducted in individuals of European descent, the limited transferability of PRS reduces its clinical value in non-European populations and may exacerbate healthcare disparities. Recent efforts to level ancestry imbalance in genomic research have expanded the scale of non-European GWAS, although most of them remain under-powered. Here we present a novel PRS construction method, PRS-CSx, which improves cross-population polygenic prediction by integrating GWAS summary statistics from multiple populations. PRS-CSx couples genetic effects across populations via a shared continuous shrinkage prior, enabling more accurate effect size estimation by sharing information between summary statistics and leveraging linkage disequilibrium (LD) diversity across discovery samples, while inheriting computational efficiency and robustness from PRS-CS. We show that PRS-CSx outperforms alternative methods across traits with a wide range of genetic architectures, cross-population genetic overlaps and discovery GWAS sample sizes in simulations, and improves the prediction of quantitative traits and schizophrenia risk in non-European populations.

## INTRODUCTION

Human complex traits and diseases do not have a single genetic cause. Instead, they are influenced by hundreds or thousands of genetic variants, each explaining a small proportion of phenotypic variation. Polygenic risk scores (PRS) aggregate genetic effects across the genome to measure the overall genetic liability to a trait or disease. PRS are not useful as a stand-alone diagnostic tool; rather, they have shown promise in predicting individualized disease risk and trajectories, stratifying patient groups, informing preventive, diagnostic and therapeutic strategies, and improving biomedical and health outcomes^1-5^. Indeed, PRS can already provide predictive power above and beyond combined clinical risk factors currently used for population screening for some diseases such as breast cancer in European populations^6,7^.

Despite the potential for clinical translation, recent theoretical and empirical studies showed that PRS have decreased cross-population prediction accuracy, especially when the discovery and target samples are genetically distant, due to a complex combination of factors including differential genetic architectures (i.e., the location of causal variants and their effect sizes), allele frequency and linkage disequilibrium (LD) patterns, phenotype definition and cohort ascertainment, and non-genetic factors and gene-by-environment interactions across populations^8-11^. As existing genome-wide association studies (GWAS) were predominantly conducted in individuals of European descent^12-15^, the poor transferability of PRS across populations has impeded its clinical implementation and raised health disparity concerns^8^. Therefore, there is an urgent need to improve the accuracy of cross-population polygenic prediction in order to maximize the clinical potential of PRS and ensure equitable delivery of precision medicine to global populations.

As the efforts to diversify the samples in genomic research start to grow, the scale of non-European genomic resources has been expanded in recent years. Although the sample sizes of most non-European GWAS remain considerably smaller than European studies, they provide critical information on the variation of genetic effects across populations. Initial studies have indicated that the genetic architectures of many complex traits and diseases are concordant between populations – both at the single-variant level and at the genome-wide level. In particular, many genetic associations identified in European populations have been replicated in non-European GWAS, and recent studies have estimated the trans-ethnic genetic correlation between European and non-European populations to be moderate or high for many traits^16-19^. These findings suggest that the transferability of PRS may be improved by integrating GWAS summary statistics from diverse populations. However, current PRS construction methods have been designed primarily for applications within one homogeneous population. Existing methods that can take GWAS summary statistics from multiple populations use meta-analysis to summarize genetic effects across training datasets^20,21^, but these approaches do not model population-specific allele frequencies and LD patterns. Alternatively, independent analysis can be performed on each discovery GWAS and the resulting PRS can be linearly combined^22,23^, but this approach does not make full use of the genetic overlap between populations to inform PRS construction. More sophisticated Bayesian polygenic prediction methods, such as LDpred^24,25^, SBayesR^26^ and PRS-CS^27^, increase prediction accuracy compared with straightforward scoring methods by improved LD modeling, but they only allow for summary statistics input from one ancestry group.

Here we present PRS-CSx, an extension of PRS-CS^27^, that improves cross-population polygenic prediction by jointly modeling GWAS summary statistics from multiple populations. PRS-CSx assumes that causal variants are largely shared across populations while allowing for their effect sizes to vary, thus leveraging the shared genetic architecture across populations to improve the accuracy of PRS while retaining modeling flexibility. In addition, PRS-CSx inherits the advantages of continuous shrinkage priors in polygenic modeling and prediction from PRS-CS, which enable accurate modeling of population-specific LD patterns and computationally efficient posterior inference. We compare the predictive performance of PRS-CSx with existing PRS construction methods across traits with a wide range of genetic architectures, cross-population genetic overlaps, and discovery GWAS sample sizes via simulations. We further apply PRS-CSx to predict quantitative traits using data from the UK Biobank (UKBB)^28^, Biobank Japan (BBJ)^29,30^, the Population Architecture using Genomics and Epidemiology Consortium (PAGE) study^31^ and the Taiwan Biobank (TWB)^32^, and predict schizophrenia risk using large-scale cohorts of European and East Asian ancestries^16,33^. We show that PRS-CSx robustly improves the prediction in non-European populations over existing methods.

## RESULTS

### Overview of PRS-CSx

PRS-CSx extends PRS-CS^27^, a recently developed Bayesian polygenic modeling and prediction framework, to improve cross-population polygenic prediction by integrating GWAS summary statistics from multiple ancestry groups (see Methods). PRS-CSx uses a shared continuous shrinkage prior to couple SNP effects across populations, which enables more accurate effect size estimation by sharing information between summary statistics and leveraging LD diversity across discovery samples. The shared prior allows for correlated but varying effect size estimates across populations, retaining the flexibility of the modeling framework. In addition, PRS-CSx explicitly models population-specific allele frequencies and LD patterns, and inherits the computational advantages of continuous shrinkage priors, and efficient and robust posterior inference algorithms (Gibbs sampling) from PRS-CS. Given GWAS summary statistics and ancestry-matched LD reference panels, PRS-CSx calculates one polygenic score for each discovery sample, and integrates them by learning an optimal linear combination to produce the final PRS (Fig. 1).

**Figure 1:**
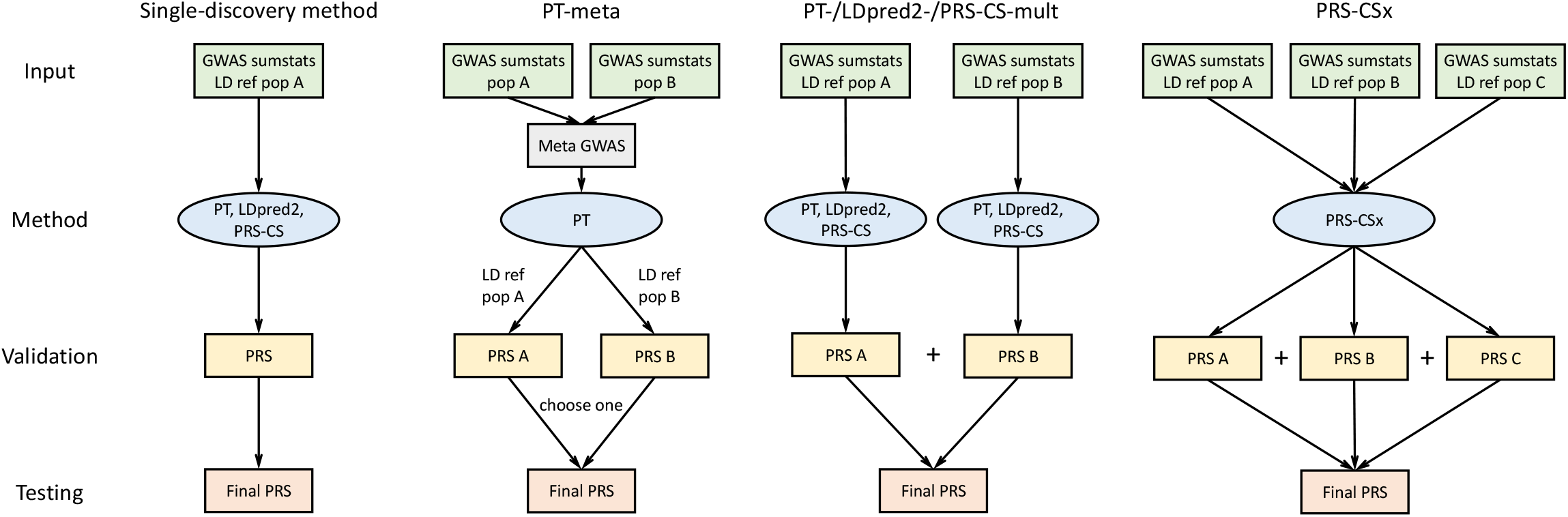
Overview of polygenic prediction methods. The predictive performances of three representative single-discovery methods: (i) LD-informed pruning and *p*-value thresholding (PT); (ii) LDpred2; (iii) PRS-CS; and five multi-discovery methods: (i) PT-meta; (ii) PT-mult; (iii) LDpred2-mult; (iv) PRS-CS-mult; (v) PRS-CSx are compared in this study. The discovery samples (to generate GWAS summary statistics), validation samples (to turn hyper-parameters in PRS construction methods) and testing samples (to assess prediction accuracy) are non-overlapping. LD ref: LD reference panel; pop A/B/C: Population A/B/C.

### Overview of PRS analysis

We have broadly classified polygenic prediction methods into two categories: *single-discovery methods*, which train PRS using GWAS summary statistics from a single discovery sample; and *multi-discovery methods*, which combines GWAS summary statistics from multiple discovery samples for PRS construction. In this work, we assess and compare within- and cross-population predictive performance of three representative single-discovery methods: (i) LD-informed pruning and *p*-value thresholding (PT)^34^; (ii) LDpred2^25^; (iii) PRS-CS^27^; and four multi-discovery methods in addition to PRS-CSx: (i) PT-meta; (ii) PT-mult^22^; (iii) LDpred2-mult; and (iv) PRS-CS-mult. PT-meta applies PT to the meta-analyzed discovery GWAS summary statistics. The three “mult” methods respectively apply PT, LDpred2 and PRS-CS to each discovery GWAS separately, and linearly combine the resulting PRS. PT-mult has been demonstrated to improve the prediction in recently admixed populations^22^. We have extended the idea of PT-mult to LDpred2-mult and PRS-CS-mult. In particular, the comparison between PRS-CSx and PRS-CS-mult can quantify the benefits of jointly modeling multiple GWAS summary statistics via the coupled shrinkage prior. The workflow for each PRS construction method is shown in Fig. 1. In all the PRS analyses, we use the *discovery* dataset to estimate the marginal effect sizes of genetic variants and generate GWAS summary statistics for each population; we use the *validation* dataset, with individual-level genotypes and phenotypes, to tune hyper-parameters for different polygenic prediction methods; and we use the *testing* dataset, with individual-level genotypes and phenotypes, to evaluate the prediction accuracy of PRS and compute performance metrics using hyper-parameters learnt in the validation dataset. The three datasets comprise non-overlapping individuals. For convenience, we use the *target* dataset to refer to the combination of *validation* and *testing* datasets, which have matched ancestry. For fair comparison across methods, we use 1000 Genomes Project (1KG) Phase 3^35^ super-population samples (European *N*=503; East Asian *N*=504; African *N*=661; Admixed American *N*=347) as the LD reference panels across different PRS construction methods throughout the paper.

### Simulations

We first evaluated the predictive performance of different polygenic prediction methods via simulations. We simulated individual-level genotypes of European (EUR), East Asian (EAS) and African (AFR) populations for HapMap3 variants with minor allele frequency (MAF) >1% in at least one of the populations using HAPGEN2^36^ and the 1KG Phase 3 samples as the reference panel. In our primary simulation setting, we randomly sampled 1% HapMap3 variants as causal variants, which in aggregation explained 50% of phenotypic variation in each population. We assumed that causal variants are shared across populations but allowed for varying effect sizes, which were sampled from a multivariate normal distribution with the cross-population genetic correlation (*r*_*g*_) set to 0.7. The simulation was repeated 20 times.

We first applied single-discovery methods (PT, LDpred2 and PRS-CS) to GWAS summary statistics generated by 100K simulated EUR samples and 20K non-EUR (EAS or AFR) samples, and evaluated their predictive performance, measured by the squared correlation (*R*^*2*^) between the simulated and predicted phenotypes, in 20K target samples, which were evenly split into a validation dataset (for hyper-parameter tuning) and a testing dataset (for the calculation of prediction accuracy) (Fig. 2; Supplementary Table 1). As expected, when the target population was EUR, PRS trained on the larger EUR GWAS were substantially more accurate than PRS trained on non-EUR GWAS (Fig. 2; left panels). However, when the target population was EAS or AFR, PRS trained on ancestry-matched non-EUR GWAS were more predictive than EUR PRS (Fig. 2; right panels), even though the sample sizes of the non-EUR GWAS were much smaller (20K vs. 100K). Among the three single-discovery methods examined, Bayesian methods (LDpred2 and PRS-CS) consistently outperformed PT. PRS-CS appeared to be more accurate than LDpred2 in both within- and cross-population prediction when the discovery GWAS was well-powered, while LDpred2 was more accurate when the discovery sample size was limited, likely reflecting the strengthens and limitations of the continuous shrinkage prior vs. the spike-and-slab prior used in PRS-CS and LDpred2, respectively. Specifically, to ensure that posterior effect size estimates are not inflated, we have imposed a minimal shrinkage to the marginal effect size estimates in PRS-CS, which may induce a stronger-than-optimal regularization when the GWAS sample size is small. In contrast, LDpred2 does not guarantee the boundedness of the posterior effect size estimates, which may better separate signals from noise when the GWAS has limited power, at the expense of the algorithm being sensitive to the imperfectly matched LD reference panel and having convergence issues when the GWAS sample size is large.

**Figure 2:**
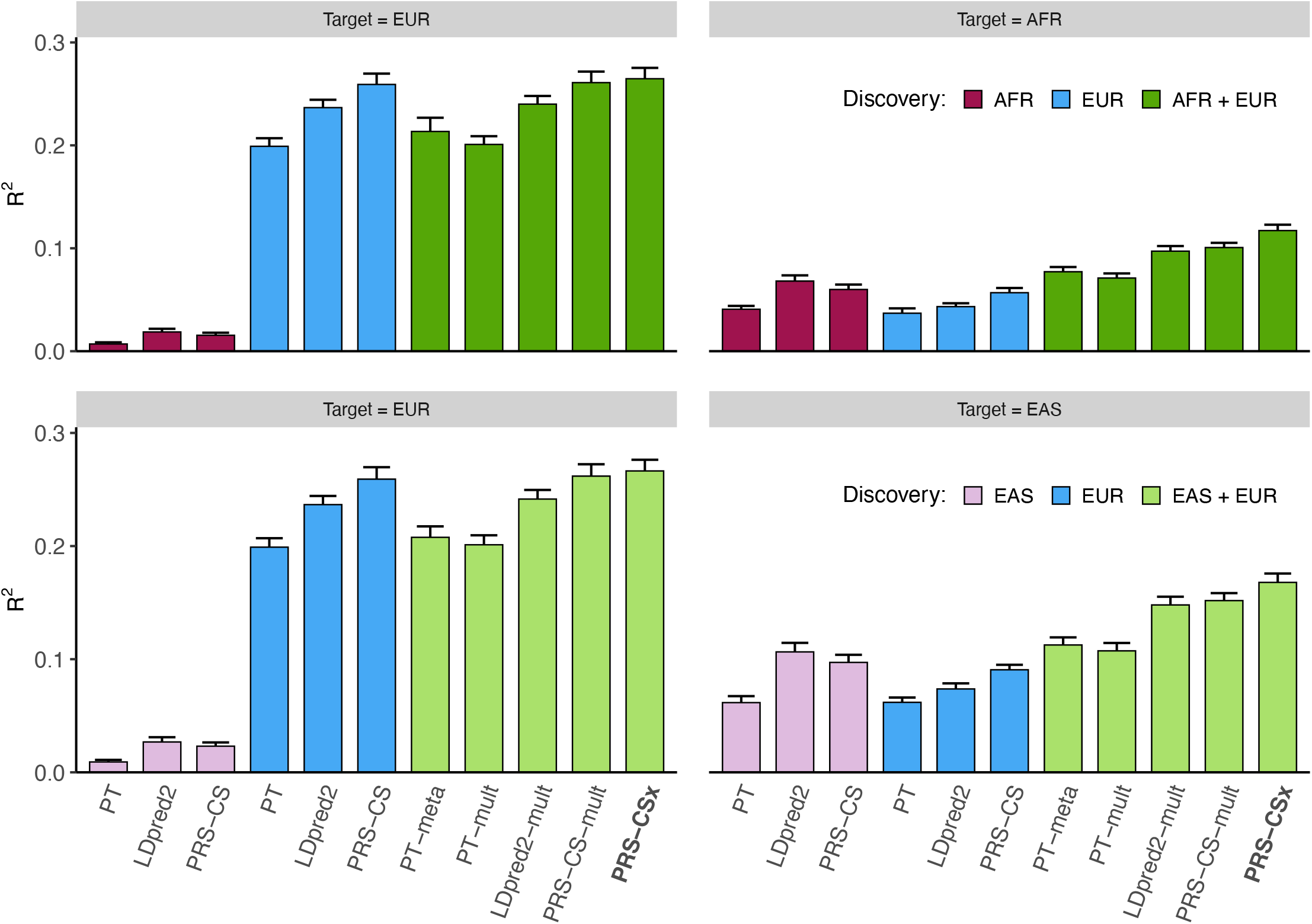
Prediction accuracy of single-discovery and multi-discovery polygenic prediction methods in simulations. 1% HapMap3 variants were randomly sampled as causal variants, which in aggregation explained 50% of phenotypic variation in each population. Causal variants were shared across populations with a cross-population genetic correlation of 0.7. 100K simulated EUR samples and 20K non-EUR (EAS or AFR) samples were used as the discovery dataset. Prediction accuracy was measured by the squared correlation (*R*^*2*^) between the simulated and predicted phenotypes in an independent testing dataset, averaged across 20 simulation replicates. Error bar indicates the standard deviation of *R*^*2*^ across simulation replicates.

We then assessed whether multi-discovery methods (PT-meta, PT-mult, LDpred2-mult, PRS-CS-mult and PRS-CSx) can improve cross-population polygenic prediction. Specifically, we used different multi-discovery methods to combine GWAS summary statistics from 100K EUR samples and 20K non-EUR (EAS or AFR) samples as the discovery dataset, and evaluated their predictive performance in independent target samples (Fig. 2; Supplementary Table 1). The smaller non-EUR GWAS relative to the EUR GWAS in this simulation design mimicked real-world scenarios where non-EUR GWAS are often underpowered. Figure 2 shows that, in general, multi-discovery methods improved prediction accuracy over their single-discovery counterparts (i.e., PT-meta or PT-mult vs. PT; LDpred2-mult vs. LDpred2; PRS-CS-mult vs. PRS-CS), reflecting the increase in discovery sample size. When the target population was EUR, the improvement of PRS-CSx and PRS-CS-mult over PRS-CS was incremental, suggesting that the benefits of adding a small non-EUR GWAS to the discovery dataset can be limited in this case. However, when predicting into non-EUR populations, Bayesian multi-discovery methods demonstrated a clear advantage over single-discovery methods. PRS-CSx provided an additional increase of 10.6% and 16.4% in *R*^2^ over PRS-CS-mult when the target population was EAS and AFR, respectively, demonstrating that joint modeling of the genetic architecture across populations using the coupled continuous shrinkage prior improves polygenic prediction in non-EUR populations. Overall, consistent with previous observations^8^, all PRS construction methods trained on EUR GWAS had decayed prediction accuracy when the genetic distance between EUR and the target populations increased^37^.

We conducted a series of secondary simulations, by varying one parameter in the primary simulation at a time, to assess the generalizability of the above observations and the robustness of PRS-CSx across a wide range of genetic architectures, cross-population genetic overlaps and discovery GWAS sample sizes. (i) We varied the polygenicity of the genetic architecture by randomly sampling 0.1% or 10% of the HapMap3 variants as causal variants. For fixed heritability, the predictive performance of all PRS construction methods reduced as the genetic architecture became more polygenic, due to the increasing difficulty of accurately estimating small genetic effects and separating signals from noise. The coupled shrinkage prior provided larger gain in prediction accuracy when the genetic architecture was sparse but was less helpful in the extreme polygenic case (Supplementary Fig. 1; Supplementary Table 2). (ii) We varied the cross-population genetic correlation using *r*_*g*_=0.4 or *r*_*g*_=1.0. As expected, cross-population prediction accuracy was higher when SNP effect sizes were highly concordant across populations and decreased when genetic effects became less correlated, making effect size estimates less transferable. However, the improvement of PRS-CSx relative to PRS-CS-mult was largely consistent across different genetic correlations (Supplementary Fig. 2; Supplementary Table 3). (iii) We varied the sample size of the discovery GWAS with the ratio of the EUR vs. non-EUR GWAS sample sizes kept unchanged (50K EUR + 10K non-EUR; 200K EUR + 40K non-EUR; 300K EUR + 60K non-EUR). PRS-CS and PRS-CS-mult were less accurate than LDpred2 and LDpred2-mult when the discovery GWAS were small but outperformed LDpred2-based methods when the GWAS became more powerful. The improvement of PRS-CSx over PRS-CS-mult was robust regardless of the variation in discovery sample size (Supplementary Fig. 3; Supplementary Table 4). (iv) We varied the ratio of the EUR vs. non-EUR GWAS sample sizes with the total sample size kept constant (120K EUR + 0K non-EUR; 80K EUR + 40K non-EUR; 60K EUR + 60K non-EUR). The prediction in non-EUR populations benefitted substantially from increasing the proportion of non-EUR training samples, and the coupled shrinkage prior provided consistent gain in prediction accuracy as the power of the non-EUR GWAS varied (Supplementary Fig. 4; Supplementary Table 5). (v) We varied the SNP heritability of the simulated trait in different populations (*h*^2^=0.5 and 0.25 in EUR and non-EUR populations respectively, and vice versa). SNP heritability determined the overall predictability of a trait, but the relative performance across different polygenic prediction methods was consistent with the primary simulation (Supplementary Fig. 5; Supplementary Table 6). (vi) We reduced the proportion of causal variants that were shared across populations to 70% or 40% to assess the robustness of PRS-CSx when the modeling assumption was violated. The transferability of PRS decreased due to reduced similarity of the genetic architecture across populations, but PRS-CSx continued to outperform alternative methods, and PRS-CS-mult in particular, suggesting that the method is robust to model misspecification (Supplementary Fig. 6; Supplementary Table 7). (vii) We simulated allele frequency and LD dependent genetic architecture, where variants with lower MAF and variants located in lower LD regions tended to have larger effects on the trait^38-40^. The predictive performance of different PRS construction methods was highly consistent with the primary simulation, suggesting that the methods we examined are not sensitive to the coupling between effect size, MAF and LD (Supplementary Fig. 7; Supplementary Table 8). (ix) Lastly, we evaluated the impact of two hyper-parameters, which determined the shape of the continuous shrinkage prior, on the predictive performance of PRS-CSx. We confirmed that the default values of the two parameters used throughout this work, which were consistent with the default setting of PRS-CS, produced optimal prediction accuracy among a grid of values we assessed (Supplementary Table 9). In summary, while the benefits of using a coupled continuous shrinkage prior varied with simulation designs and may be small in certain scenarios, we concluded that PRS-CSx improved cross-population prediction accuracy relative to alternative methods across a vast majority of the simulation settings and was robust to model misspecification.

### Prediction of quantitative traits in Biobanks

Next, we evaluated the predictive performance of different polygenic prediction methods using 33 anthropometric or blood panel traits from UKBB^28^ (*N*=314,916-360,388) and BBJ^30^ (*N*=71,221-165,419; Supplementary Table 10). Summary statistics were acquired from the Neale Lab (UKBB GWAS) and BBJ website (Data availability). All the 33 traits, with two exceptions (Basophil and Eosinophil), had moderate to high cross-population genetic-effect correlations estimated by POPCORN^17^ (range 0.37-0.85; Supplementary Table 10). We applied single-discovery methods to UKBB or BBJ summary statistics, and used multi-discovery methods to combine UKBB and BBJ GWAS. All target samples are unrelated UKBB individuals that are also unrelated with the UKBB discovery samples. We assigned each target sample to one of the five 1KG super-populations [AFR, AMR (Admixed American), EAS, EUR, SAS (South Asian)] (see Methods), and assessed the prediction accuracy in each target population separately, adjusting for age, sex and top 20 principal components (PCs) of the genotypes. For each population, the target dataset was randomly and evenly split into a validation dataset (for hyper-parameter tuning) and a testing dataset (for the evaluation of predictive performance). The prediction accuracy, measured by variance explained (*R*^*2*^) in linear regression after adjusting for covariates, was averaged across 100 random splits.

Consistent with simulation results, Bayesian multi-discovery methods (LDpred2-mult, PRS-CS-mult and PRS-CSx) often outperformed single-discovery methods and PT-based multi-discovery methods, suggesting the importance of integrating available GWAS summary statistics and appropriately accounting for population-specific LD patterns in cross-population prediction (Fig. 3a; Supplementary Table 11). The improvement of PRS-CSx in prediction accuracy relative to LDpred2 and PRS-CS trained on UKBB summary statistics (which on average were more accurate than PRS trained on BBJ summary statistics), and LDpred2-mult and PRS-CS-mult (which were often the second and third best multi-discovery method) depended on the target population.

**Figure 3:**
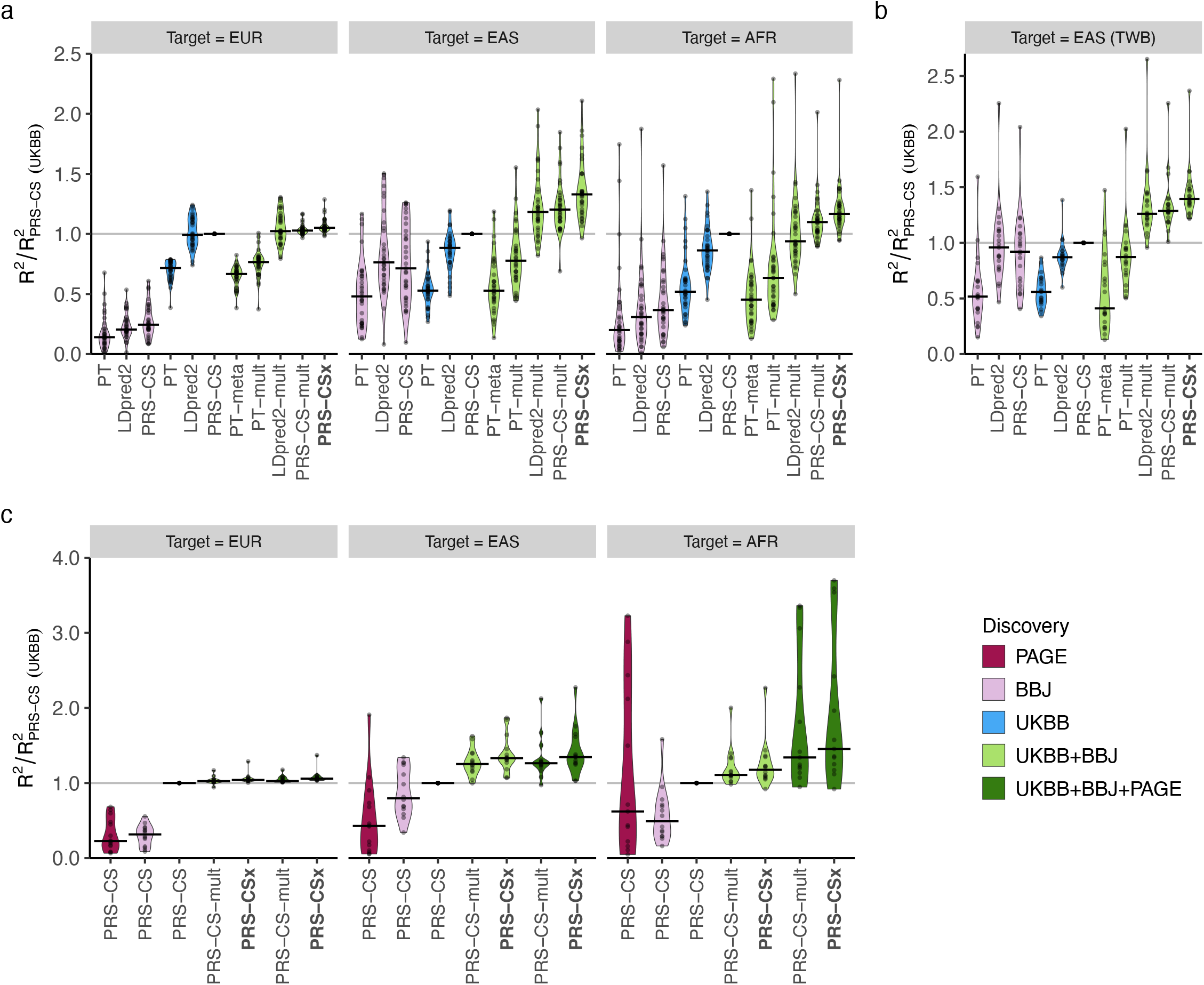
Prediction accuracy of quantitative traits using GWAS summary statistics from UKBB, BBJ and PAGE. (a) Relative prediction accuracy of single-discovery and multi-discovery polygenic construction methods using GWAS summary statistics from UKBB and BBJ, with respect to PRS-CS trained on the UKBB GWAS, across 33 traits in different UKBB target populations (EUR, EAS and AFR). Each data point shows the relative prediction *R*^*2*^ of a trait, averaged across 100 random splits of the target samples into validation and testing datasets. (b) Relative prediction accuracy of different polygenic prediction methods using discovery GWAS from UKBB and BBJ, with respect to PRS-CS trained on the UKBB GWAS, across 21 traits in the Taiwan Biobank (TWB). Each data point shows the relative prediction *R*^*2*^ of a trait. (c) Relative prediction accuracy of PRS-CS, PRS-CS-mult and PRS-CSx using discovery GWAS from UKBB, BBJ and PAGE, with respect to PRS-CS trained on the UKBB GWAS, across 14 traits in different UKBB target populations (EUR, EAS and AFR). Each data point shows the relative prediction *R*^*2*^ of a trait, averaged across 100 random splits of the target samples into validation and testing datasets. MCHC was removed from the AFR panel due to the near-zero *R*^*2*^ of PRS-CS (UKBB) and thus inflated relative prediction *R*^*2*^ for other methods. In all panels, the crossbar indicates the median of relative *R*^*2*^ across traits examined.

When predicting into the EUR population, PRS-CSx provided a consistent but marginal improvement over LDpred2 (median relative increase in *R*^2^: 4.7%) and PRS-CS (median relative increase in *R*^2^: 5.2%), likely due to the limited power of the BBJ GWAS relative to the UKBB GWAS. The benefit of the coupled prior in this case was also limited, as reflected by a small improvement of PRS-CSx relative to PRS-CS-mult (median relative increase in *R*^2^: 2.2%; Fig. 3a, left panel; Supplementary Table 11), which was consistent with the observations in simulations. When the target population was EAS, however, PRS-CSx substantially increased the prediction accuracy relative to single-discovery methods: the median relative improvements in *R*^*2*^ were 52.3% and 32.9% when compared with LDpred2 and PRS-CS trained on UKBB GWAS, and 69.8% and 74.4% when compared with LDpred2 and PRS-CS trained on BBJ GWAS, suggesting that PRS-CSx can leverage large-scale EUR GWAS to improve the prediction in non-EUR populations. PRS-CSx also had a median improvement of 10.5% and 8.3% relative to LDpred2-mult and PRS-CS-mult, respectively, demonstrating the benefits of jointly modeling summary statistics from multiple populations in trans-ancestry prediction (Fig. 3a, middle panel; Supplementary Table 11). When the target population did not match any of the discovery samples, PRS-CSx was still able to boost the prediction accuracy. For example, when predicting into the AFR population, the median improvements of PRS-CSx relative to UKBB LDpred2 and PRS-CS were 45.1% and 16.9%, respectively, and the median improvements relative to LDpred2-mult and PRS-CS-mult were 22.2% and 7.1%, respectively (Fig. 3a, right panel; Supplementary Table 11). That said, the prediction accuracy in the AFR population remained low relative to the predictions in EUR and EAS populations, probably because both discovery samples are genetically distant.

We next sought to replicate the relative performance of different PRS construction methods in the Taiwan Biobank (TWB)^32^, which is a community-based prospective cohort study of the Taiwanese population. Among the 33 quantitative traits we examined in UKBB and BBJ, 21 traits were also available in TWB. All PRS were trained on the UKBB and/or BBJ GWAS, validated in the UKBB EAS samples (where hyper-parameters were learnt), and evaluated in the TWB sample comprising 10,149 unrelated individuals, adjusting for age, sex and top 20 PCs of the genotypes. Figure 3b shows that single-discovery methods trained on UKBB GWAS and BBJ GWAS had similar performance in the TWB sample, even though UKBB GWAS were much larger (Fig. 3b; Supplementary Table 12). Bayesian multi-discovery methods showed substantial improvement in prediction accuracy compared with single-discovery methods. PRS-CSx provided a median improvement of 39.5% relative to PRS-CS (best single-discovery method) and 8.2% relative to PRS-CS-mult (second best multi-discovery method), suggesting the robustness of PRS-CSx when model parameters learnt in validation datasets were applied to external independent testing datasets. Overall, results in the TWB closely reproduced the patterns observed in the UKBB EAS samples (Fig. 3a, middle panel).

We further investigated whether adding African American samples to the discovery dataset can improve the prediction in the AFR population. Among the 33 traits we examined, 14 traits were also available in PAGE^31^ (*N*=11,178-49,796), a genetic epidemiology study comprising largely of African American and Hispanic/Latino samples (Supplementary Table 10). All UKBB-PAGE and BBJ-PAGE genetic-effect correlations were moderate to high (range 0.44-1.00; Supplementary Table 10). Although the discovery and target samples had largely matched ancestry, applying PRS-CS (or other single-discovery methods) to PAGE summary statistics alone produced low prediction accuracy in the AFR population with only a few exceptions, due to the small sample size of the PAGE study (Fig. 3c; Supplementary Table 13). However, integrating UKBB, BBJ and PAGE summary statistics using PRS-CSx dramatically outperformed single-discovery methods, and the median relative improvement in *R*^*2*^ was 28.1% when compared with PRS-CSx trained on UKBB and BBJ GWAS only, suggesting that PRS-CSx benefits from including samples that have matched ancestry with the target population in the discovery dataset, even if the non-European GWAS included are considerably smaller than European studies (Fig. 3c; Supplementary Table 13).

### Schizophrenia risk prediction

Lastly, we evaluated the predictive performance of different polygenic prediction methods for dichotomous traits. We used schizophrenia as an example, for which large-scale EUR and EAS samples are available (Supplementary Table 14). Specifically, we used GWAS summary statistics derived from the Psychiatric Genomics Consortium (PGC) wave 2 EUR samples (33,640 cases and 43,456 controls)^33^ and 10 PGC EAS cohorts^16^ (7,856 cases and 11,562 controls) as the discovery dataset. For the additional 7 EAS cohorts which we had access to individual-level data, we set aside one cohort (687 cases and 492 controls) as the validation dataset (for hyper-parameter tuning), and applied a leave-one-out approach to the remaining 6 cohorts. More specifically, we in turn used one of the 6 cohorts as the testing dataset, and meta-analyzed the remaining 5 cohorts with the 10 PGC EAS cohorts using an inverse-variance-weighted meta-analysis to generate the discovery GWAS summary statistics for the EAS population. The prediction accuracy of different PRS construction methods was then evaluated in the left-out (testing) cohort, adjusting for sex and top 20 PCs. As a reference, the SNP heritability of schizophrenia on the liability scale was estimated to be 0.23±0.03 in the EAS population^16^.

Consistent with previous observations, PRS trained on EAS GWAS were more predictive in EAS cohorts than those trained on PGC EUR summary statistics^16^, despite the larger sample size for the EUR GWAS (Fig. 4a; Supplementary Table 15). Among single-discovery methods, LDpred2 and PRS-CS performed substantially better than PT, highlighting the importance of modeling LD patterns for highly polygenic traits. By integrating EUR and EAS summary statistics, Bayesian multi-discovery methods dramatically increased the prediction accuracy relative to single-discovery methods. In particular, compared with LDpred2, the best-performing single-discovery method in this analysis, PRS-CSx increased the median *R*^*2*^ on the liability scale (assuming 1% of disease prevalence) from 0.043 (LDpred2 trained on EAS GWAS) and 0.031 (LDpred2 trained on EUR GWAS) to 0.063, a relative increase of 45.4% and 104.9%, respectively. PRS-CSx also approximately doubled the prediction accuracy of PT-meta and PT-mult in the median liability *R*^*2*^, with a relative increase of 135.9% (from 0.027 to 0.063) and 95.3% (from 0.032 to 0.063), respectively. In addition, PRS-CSx provided consistent, although relatively small, improvement over LDpred2-mult (relative increase in median *R*^*2*^: 8.7%) and PRS-CS-mult (relative increase in median *R*^*2*^: 5.9%), suggesting that the coupled prior can be useful in practice even for highly polygenic architecture (Fig. 4a; Supplementary Table 15). Other performance metrics, including Nagelkerke’s *R*^*2*^, odds ratio (OR) per standard deviation change of PRS, and OR comparing top 10% with bottom 10% of the PRS distribution, showed a consistent pattern (Supplementary Table 15). Finally, PRS-CSx can more accurately identify individuals at high/low schizophrenia risk than alternative methods, showing a 2.9, 3.5 and 4.2-fold increase in case prevalence across the 6 testing cohorts when contrasting the top 10%, 5% or 2% of the PRS distribution with the bottom 10%, 5% or 2%, respectively (Fig. 4b; Supplementary Table 16).

**Figure 4:**
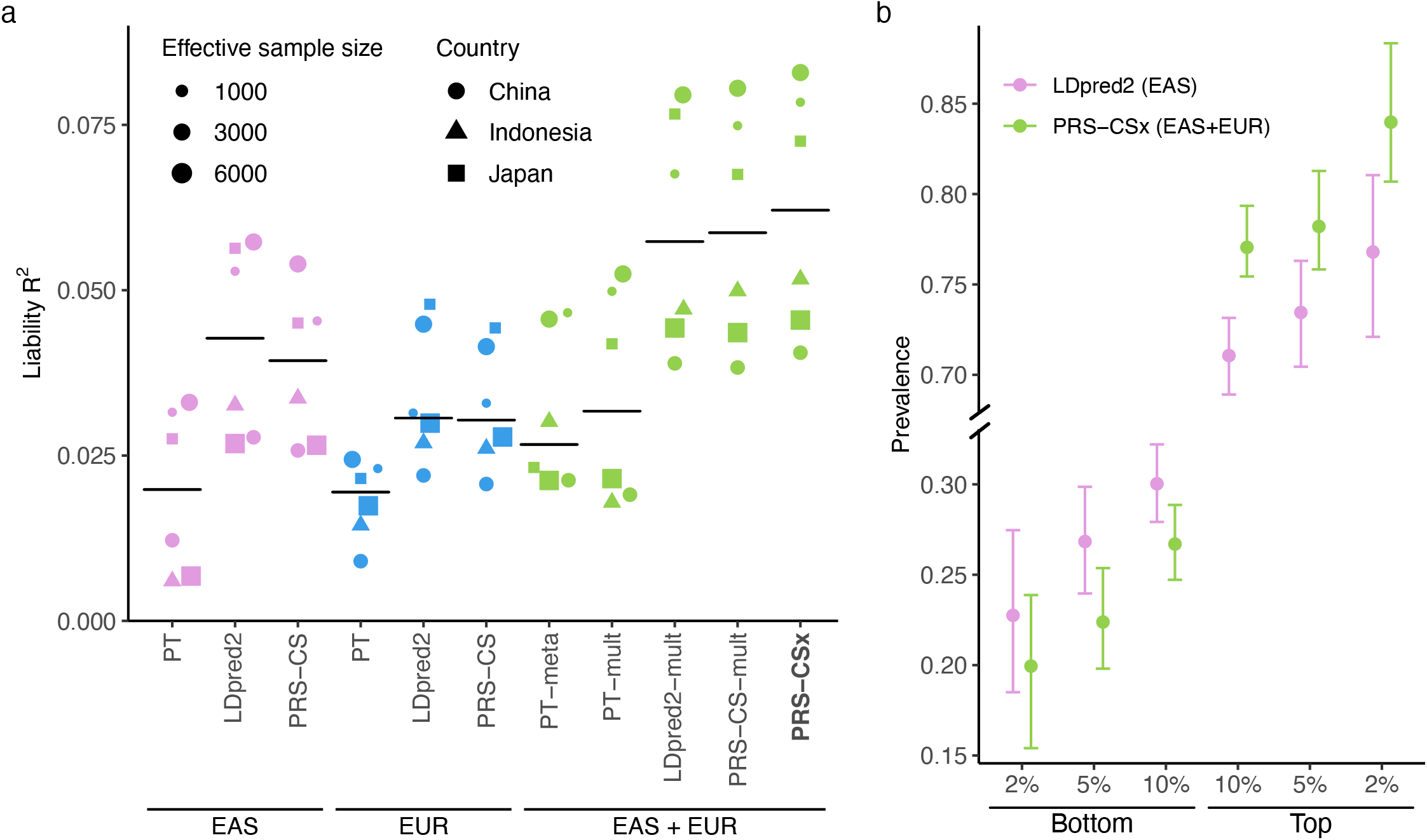
Prediction accuracy of schizophrenia risk in EAS cohorts. (a) Prediction accuracy, measured as *R*^*2*^ on the liability scale, of single-discovery (trained on EAS or EUR GWAS) and multi-discovery polygenic prediction methods (trained on both EAS and EUR GWAS: EAS+EUR) across 6 EAS schizophrenia cohorts. Each dot represents one testing cohort, with the size of the dot proportional to the effective sample size, calculated as 4(1/Ncase+1/Ncontrol), and the shape of the dot representing the country where the sample was collected. Crossbar indicates the median *R*^*2*^ on the liability scale. (b) Case prevalence of the bottom 2%, 5%, 10% and top 2%, 5%, 10% of the PRS distribution, constructed by LDpred2 trained on EAS GWAS (the best-performing single-discovery method) and PRS-CSx (the best-performing multi-discovery method), across 6 EAS schizophrenia cohorts. Error bar indicates 95% confidence intervals.

## DISCUSSION

We have presented PRS-CSx, a Bayesian polygenic prediction method that integrates GWAS summary statistics from multiple populations to improve the prediction accuracy of PRS in ancestrally diverse samples. PRS-CSx leverages the correlation of genetic effects and LD diversity across populations to more accurately localize association signals and increase the effective sample size of the discovery dataset, while accounting for population-specific allele frequency and LD patterns. We have shown, via simulation studies, that PRS-CSx robustly improves cross-population prediction over existing methods across traits with varying genetic architectures, genetic overlaps between populations, and discovery GWAS sample sizes (Fig. 2). Using quantitative traits from multiple biobanks (Fig. 3) as well as schizophrenia cohort studies of European and East Asian ancestries (Fig. 4), we have further demonstrated the PRS-CSx can leverage large-scale European GWAS to boost the accuracy of polygenic prediction in non-European populations, for which ancestry-matched discovery GWAS may be orders of magnitude smaller in sample size.

PRS-CSx is designed to flexibly model GWAS summary statistics from multiple populations where SNP effect sizes and/or LD patterns differ. For two or more GWAS conducted in independent samples from the same population where effect sizes and LD patterns are expected to be highly concordant, a fixed-effect meta-analysis, whose modeling assumptions are fully satisfied in this scenario, is probably the optimal approach to combine the GWAS and maximize statistical power. PRS-CSx appears to be most effective when one of the discovery samples has matched ancestry with the target population, although it may be able to boost prediction in populations distant from any of the training samples as well. While PRS-CSx increased the prediction for the majority of the traits we examined, the amount of improvement in prediction accuracy over alternative methods varied across traits and may depend on multiple factors including the comparative genetic architecture across populations, the power of the discovery GWAS and the sample characteristics of the target dataset. Future research is needed to dissect the effects of potential factors on the accuracy of cross-ancestry polygenic prediction and to better understand the behavior of different prediction algorithms for individual traits.

Convergence of the MCMC samplers employed by Bayesian polygenic prediction methods is often overlooked in the literature, likely because (i) the focus of polygenic prediction is to aggregate genetic effects across the genome into a single score, rather than making inference of the genetic effects of individual variants; and (ii) the sheer size of the model parameters (>1 million for each population) makes traditional model diagnostic methods, which often rely on graphical outputs, difficult to apply. To assess the overall convergence of the Gibbs sampler used in PRS-CSx, for each trait, we selected a few SNPs where we monitored the convergence of their posterior effect size estimates. Some of the SNPs had strong associations with the trait in multiple populations, while some of the SNPs were null across populations. We ran the PRS-CSx model three times using different random seeds, and assessed the convergence using the Gelman-Rubin convergence diagnostic for multiple chains^41^. All reduction factors across the SNPs we examined were smaller than 1.05, indicating convergence. As an example, Supplementary Fig. 8 shows the trace plots and autocorrelation functions (ACFs) for the posterior effects of rs7412 on low-density lipoprotein cholesterol (LDL-C) when integrating UKBB, BBJ and PAGE GWAS summary statistics using PRS-CSx. This SNP, located within the *APOE* locus on chromosome 19, had extremely strong marginal associations with LDL-C across the three populations (all *P*-values <1E-200). Trace plots and ACFs indicated that the Gibbs sampler achieved reasonable convergence and mixing. Future work is needed to better monitor the behavior of the Markov chain in high-dimensional settings and characterize the uncertainties of the PRS constructed by Bayesian methods due to the randomness of the posterior sampler^42^.

While PRS-CSx can take an arbitrary number of GWAS summary statistics as input, an ancestry-matched LD reference panel is required for each discovery sample, which may be challenging to build for GWAS conducted in admixed populations or in samples with large genomic diversity. For example, in this work, we have used a reference panel built from the 1KG AMR samples to approximate the LD patterns in the PAGE GWAS, which largely comprised of African American and Hispanic/Latino samples^31^. Although we have shown that this approach improved the prediction in the African population, indicating that PRS-CSx may be robust to imperfectly matched LD reference panels, which is also consistent with recent independent evaluations^23^, we note that this is a panel built from a small number of individuals that may not accurately capture the LD patterns in the discovery sample. Future work is needed to better model summary statistics from recently admixed populations. One promising direction is to characterize and incorporate local ancestries into PRS construction using ancestry-specific effect size estimates^43,44^. The Bayesian modeling framework of PRS-CSx and the flexibility of continuous shrinkage priors also allow for the incorporation of functional annotations and fine-mapping results into PRS construction to improve the portability of PRS, as shown by recent studies^23,45^. Moreover, given that many diseases, such as autoimmune diseases and psychiatric disorders, have substantial genetic overlap, extending the PRS-CSx framework to integrate genetically correlated traits may further inform the construction of PRS and increase their prediction accuracy^46^.

Lastly, we note that although PRS-CSx can improve cross-population polygenic prediction, the gap in prediction accuracy between European and non-European populations, especially the African population, remains considerable, and many predictions in non-European populations are not practically useful. Indeed, sophisticated statistical and computational methods alone will not be able to overcome the current Eurocentric biases in GWAS. Broadening the sample diversity in genomic research to fully characterize the genetic architecture and understand the genetic and non-genetic contributions to human complex traits and diseases across global populations is crucial to further improve the prediction accuracy of PRS in diverse populations. We believe that the rapid expansion of genomic resources for non-European populations coupled with advanced analytic methods will accelerate the equitable deployment of PRS in clinical settings and maximize its healthcare potential.

## Data Availability

Publicly available data were downloaded from the following databases: 1000 Genomes Project Phase 3 reference panels: https://mathgen.stats.ox.ac.uk/impute/1000GP_Phase3.html; Genetic map for each subpopulation: ftp.1000genomes.ebi.ac.uk/vol1/ftp/technical/working/20130507_omni_recombination_rates; UKBB summary statistics: http://www.nealelab.is/uk-biobank ("GWAS round 2" was used in this study); BBJ summary statistics were downloaded from PheWeb: https://pheweb.jp; PAGE summary statistics were downloaded from the GWAS Catalog: https://www.ebi.ac.uk/gwas/downloads/summary-statistics; PGC wave 2 schizophrenia GWAS (49 EUR samples): https://www.med.unc.edu/pgc/download-results/; Schizophrenia EAS summary statistics are available upon request to the PGC Schizophrenia Work Group. Individual-level schizophrenia data of East Asian ancestry are available upon reasonable request with regulatory/compliance review to the Stanley Global Asia Initiatives.

## AUTHOR CONTRIBUTIONS

H.H. and T.G. designed the project; T.G. developed the statistical methods and programmed the code for PRS-CSx; Y.R. and T.G. conducted simulation studies; Y.R. and T.G performed the analysis in the UK Biobank; Y.F.L. performed the analysis in the Taiwan Biobank; Y.R. performed the analysis in the schizophrenia cohorts; Y.A.F. assigned the UKBB samples into super-population groups; C.C. provided critical suggestions for the study design; M.L. took part in the testing of the code and preprocessed schizophrenia East Asian data; Z.G., L.H., A.S. and S.Q. contributed to the generation and preprocessing of schizophrenia East Asian data. Y.R., H.H. and T.G. wrote the manuscript; Y.A.F. and A.R.M. provided critical revision for the manuscript; All the authors reviewed and approved the final version of the manuscript.

## COMPETING INTERESTS

C.C. is an employee of Biogen. The other authors declare no competing interests.

## ACKNOWLEDGEMENTS

We thank the Neale Lab and Biobank Japan (BBJ) for releasing the genome-wide association summary statistics from UK Biobank (UKBB) and BBJ. Individual-level phenotypes and genotypes for UKBB samples were obtained under application 32568. We thank the Psychiatric Genomics Consortium (PGC) for providing the GWAS summary statistics for schizophrenia. T.G. is supported by NIA K99/R00AG054573. H.H. acknowledges support from NIDDK K01DK114379, NIMH U01MH109539, Brain & Behavior Research Foundation Young Investigator Grant, the Zhengxu and Ying He Foundation, and the Stanley Center for Psychiatric Research. S.Q. is supported by Shanghai Municipal Science and Technology Major Project (Grant No. 2017SHZDZX01). A.R.M. is supported by NIMH K99/R00MH117229.

## CODE AVAILABILITY

The code used in this study is available from the following websites: PRS-CSx: https://github.com/getian107/PRScsx; PRS-CS: https://github.com/getian107/PRScs; LDpred2: https://privefl.github.io/bigsnpr/articles/LDpred2; PRSice2: https://www.prsice.info; HAPGEN2: https://mathgen.stats.ox.ac.uk/genetics_software/hapgen/hapgen2.html; PLINK: https://www.cog-genomics.org/plink/; LD score regression: https://github.com/bulik/ldsc; POPCORN: https://github.com/brielin/Popcorn; Interpolation of genetic maps: https://github.com/joepickrell/1000-genomes-genetic-maps; Population assignment: https://github.com/Annefeng/PBK-QC-pipeline.

## METHODS

### PRS-CSx

PRS-CSx is an extension of PRS-CS^27^, which enables the integration of GWAS summary statistics from multiple populations to improve cross-population polygenic prediction. Consider the following Bayesian linear regression for *K* populations:

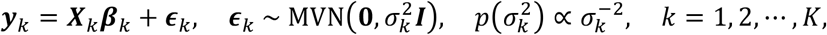

where, for each population *k*, ***y***_*k*_ is a vector of standardized phenotypes (zero mean and unit variance) from *N*_*k*_ individuals, ***X***_*k*_ is an *N*_*k*_ ×*M* matrix of standardized genotypes (each column has zero mean and unit variance), ***β***_*k*_ is a vector of SNP effect sizes, ***ϵ***_*k*_ is a vector of normally distributed non-genetic effects with variance 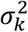, for which we assign a non-informative scale-invariant Jeffreys prior, and ***I*** is an identify matrix. For SNP *j* in population *k*, we place a continuous shrinkage prior on its effect size *β*_*jk*_, which can be represented as global-local scale mixtures of normals:

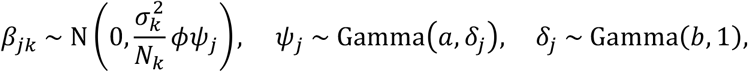

where *ϕ* is a global shrinkage parameter shared across all SNPs that models the overall sparseness of the genetic architecture, and *ψ*_*j*_ is a local, SNP-specific shrinkage parameter that is adaptive to marginal GWAS associations. By assigning a gamma-gamma hierarchical prior on *ψ*_*j*_ (specifically, the Strawderman-Berger prior with *a* = 1 and *b* = 1/2 in this work), the marginal prior density of *β*_*jk*_ has sizable amount of mass near zero to impose strong shrinkage on small noisy signals, and in the meantime, heavy Cauchy-like tails to avoid over-shrinkage of truly non-zero effects.

We note that when SNP *j* is available in multiple GWAS summary statistics, the continuous shrinkage prior is shared across populations (i.e., both *ϕ* and *ψ*_*j*_ do not depend on *k*), enabling information sharing between summary statistics while allowing for varying SNP effect sizes across populations to retain modeling flexibility. More specifically, given the variance parameters 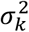, *ϕ* and *ψ*_*j*_, and the marginal least squares estimates of the SNP effect sizes in population 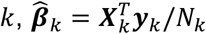, the posterior mean of ***β***_*k*_ is 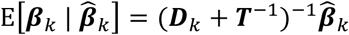, where 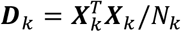 is the LD matrix for population *k*, and ***T*** = diag{*ϕψ*_1_, *ϕψ*_2_, ⋯, *ϕψ*_*M*_} is a diagonal matrix containing both global and local shrinkage parameters. It can be seen that ***T*** does not depend on *k* and thus the amount of shrinkage applied to each SNP is shared across populations. Meanwhile, population-specific LD patterns are explicitly modelled via the LD matrix ***D***_*k*_.

Given the summary statistics and ancestry-matched LD reference panel for each discovery sample, the PRS-CSx model can be fitted using a Gibbs sampler with block update of posterior SNP effect sizes, without the need to access individual-level data (Supplementary Methods). Monomorphic or rare variants not present in the population-specific LD reference panel of population A are not included in the construction of PRS for population A. If a SNP is present in population A but is monomorphic or rare in other populations, its effect size is not coupled in posterior inference but the SNP is included in the PRS of population A such that population-specific associations can be captured (Fig. 1). PRS-CSx inherits many features from PRS-CS, including robustness to varying genetic architectures, multivariate modeling of population-specific LD patterns, and computational efficiency. In this work, we used pre-calculated 1KG Phase 3 LD reference panels^47^ for EUR, EAS, AFR and AMR populations, which were constructed for HapMap3 variants with MAF >1%. We recommend using 1,000\**K* Markov Chain Monte Carlo (MCMC) iterations with the first 500\**K* steps as burin-in in Gibbs sampling, where *K* is the number of discovery populations, reflecting the growing number of unknown parameters with the number of discovery GWAS jointly modelled. For a fixed global shrinkage parameter *ϕ*, PRS-CSx returns posterior SNP effect size estimates for each discovery population, which can be used to calculate PRS in the target sample. For each *ϕ* value, we fitted a linear regression of the z-scored PRS (one for each discovery population) in the validation dataset:

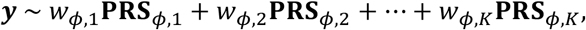

where ***y*** is the trait of interest, **PRS**_*ϕ, k*_ is the standardized PRS for population *k*, and *w*_*ϕ, k*_ is the regression coefficient. We screened four different *ϕ* values, 10^−6^, 10^−4^, 10^−2^ and 1.0, in this work. The *ϕ* value and the corresponding regression coefficients for the linear combination of PRS that maximized the *R*^*2*^ in the validation dataset were used in the testing dataset to calculate the final PRS:

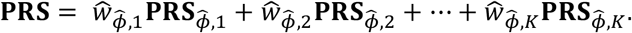

### Existing PRS methods

#### PT

LD-informed pruning and *p*-value thresholding (PT)^34^ selects clumped SNPs of a certain statistical significance to be included in the PRS calculation. We performed PT using PRSice-2^48^ with the default parameter settings: the clumping was performed with a radius of 250kb and an *r*^*2*^ threshold of 0.1. We used 1KG super-population samples (EUR, EAS, AFR or AMR) whose ancestry matched the discovery sample as the LD reference panel for clumping. The *p*-value threshold among 10^−8^, 10^−7^, 10^−6^, 10^−5^, 3×10^−5^, 10^−4^, 3×10^−4^, 0.001, 0.003, 0.01, 0.03, 0.1, 0.3 and 1.0 that maximized the *R*^*2*^ in the validation dataset was selected, and used in the independent testing dataset to calculate the final PRS and its performance metrics.

#### LDpred2

LDpred2^25^, an improved version of the LDpred algorithm^24^, is a Bayesian polygenic prediction method that adjusts marginal SNP effect size estimates from GWAS summary statistics to calculate the PRS. LDpred2 assigns a point-normal prior to SNP effect sizes, where the proportion of causal variants is a tunable parameter, and infers posterior effects using a Gibbs sampler. We constrained the computation to HapMap3 variants with MAF >1%, and used 1KG super-population samples (EUR, EAS, AFR or AMR) whose ancestry matched the discovery sample as the LD reference panel. We ran LDpred2-grid using the genome-wide option with the full LD matrix, and tested the proportion of causal variants from a sequence of 17 values equally spaced from 10^−4^ to 1.0 on the log scale. The proportion that maximized the *R*^*2*^ in the validation dataset was selected, and used in the independent testing dataset to calculate the final PRS and its performance metrics.

#### PRS-CS

PRS-CS^27^ is a Bayesian polygenic prediction method that infers posterior SNP effect sizes from summary statistics using a continuous shrinkage prior, which is robust to varying genetic architectures, accurate in LD modeling and computationally efficient. PRS-CS has one hyper-parameter, the global shrinkage parameter, which models the overall sparseness of the genetic architecture. We used default parameter settings and the pre-calculated 1KG LD reference panel (EUR, EAS, AFR or AMR) that matched the ancestry of the discovery sample, which was constructed for HapMap3 variants with MAF >1%. The global shrinkage parameter among 10^−6^, 10^−4^, 10^−2^ and 1.0 that maximized the *R*^*2*^ in the validation dataset was selected, and used in the independent testing dataset to calculate the final PRS and its performance metrics.

#### PT-meta

PT-meta applies PT to the meta-GWAS that combines all discovery summary statistics through an inverse-variance-weighted fixed-effect meta-analysis. We used the same clumping parameters and screened the same list of *p*-value thresholds as the PT method. The 1KG LD reference panel (EUR, EAS, AFR or AMR) that had matched ancestry with each of the discovery samples was in turn used for clumping, producing multiple sets of clumped variants. The best combination of the LD reference panel and the *p*-value threshold that maximized the *R*^*2*^ in the validation dataset was selected, and used in the independent testing dataset to calculate the final PRS and its performance metrics.

#### PT-mult, LDpred2-mult and PRS-CS-mult

PT-mult^22^, LDpred2-mult and PRS-CS-mult apply PT, LDpred2 and PRS-CS to each discovery summary statistics separately. The most predictive PRS derived from each discovery sample were then used to fit a linear regression in the validation dataset:

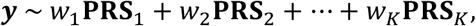

where **PRS**_*k*_ is the standardized PRS for population *k*, and *w*_*k*_ is the corresponding regression coefficient. The optimal hyper-parameter for each discovery sample and the estimated regression coefficients for the linear combination of standardized PRS were used in the independent testing dataset to calculate the final PRS and its performance metrics. We screened the same grid of hyper-parameters for each method (i.e., the clumping parameter and *p*-value threshold for PT; the proportion of causal variants for LDpred2; and the global shrinkage parameter for PRS-CS). The 1KG super-population samples (EUR, EAS, AFR or AMR) whose ancestry matched the discovery sample were used as the LD reference panel.

### Simulations

#### Genotypes

We simulated individual-level genotypes of EUR, EAS and AFR populations using HAPGEN2^36^ and ancestry-matched 1KG Phase 3^35^ super-population samples as the reference panel. We grouped CEU, IBS, FIN, GBR and TSI into the EUR super-population, CDX, CHB, CHS, JPT and KHV into the EAS super-population, and ACB, ASW, LWK, MKK and YRI into the AFR super-population. To calculate the genetic map (cM) and recombination rate (cM/Mb) for each super-population, we downloaded the maps and rates for their constituent subpopulations (Data availability), linearly interpolated the genetic map and recombination rate at each position (Code availability), and averaged the genetic maps and recombination rates across the subpopulations within each super-population. We simulated 320K EUR samples, 100K EAS samples and 100K AFR samples, and confirmed that the allele frequencies and LD patterns of the simulated genotypes were highly similar to those of the 1KG reference panels. 20K samples for each of the three populations were saved as the target dataset, which was evenly split into validation and testing datasets. The remaining samples served as the discovery dataset, which was used to produce GWAS of varying sample sizes. We constrained the simulations to 1,296,253 HapMap3 variants with MAF >1% in at least one of the EUR, EAS and AFR populations, and removed triallelic and strand ambiguous variants.

#### Phenotypes

In our primary simulation, we randomly sampled 1% of the HapMap3 variants as causal variants. We assumed that causal variants are shared across the three populations and simulated their per-allele effect sizes using a multivariate normal distribution with the correlation between populations set to 0.7. For each population, we used a normally distributed random variable to model the non-genetic component such that the heritability was fixed at 50%. The phenotype was then generated in each population using ***y*** = ***Xβ*** + ***ϵ***, where ***X*** was the genotype matrix, ***β*** was the simulated per-allele effect size vector in which causal variants had non-zero effects and the rest of the variants had zero effect sizes, and ***ϵ*** was the simulated non-genetic component. The simulation was repeated 20 times. GWAS was performed on 100K EUR, 20K EAS and 20K AFR discovery samples, respectively, using PLINK 1.9^49^, to generate summary statistics.

We conducted a series of secondary simulations to assess the robustness of PRS-CSx in a wide range of settings: (i) varying polygenicity of the genetic architecture (0.1% vs. 1% vs. 10% of causal variants); (ii) varying cross-population genetic correlations (*r*_*g*_=0.4 vs. *r*_*g*_=0.7 vs. *r*_*g*_=1.0); (iii) varying sample sizes of the discovery GWAS (50K EUR + 10K non-EUR; 100K EUR + 20K non-EUR; 200K EUR + 40K non-EUR; 300K EUR + 60K non-EUR); (iv) varying ratios of the EUR vs. non-EUR GWAS sample sizes (120K EUR + 0K non-EUR; 100K EUR vs. 20K non-EUR; 80K EUR + 40K non-EUR; 60K EUR + 60K non-EUR); (v) varying SNP heritability of the simulated trait in different populations (*h*^2^=0.5 in EUR + *h*^2^=0.5 in non-EUR; *h*^2^=0.5 in EUR + *h*^2^=0.25 in non-EUR; *h*^2^=0.25 in EUR + *h*^2^=0.5 in non-EUR); (vi) varying proportions of shared causal variants across populations (100% vs. 70% vs. 40%); (vii) allele frequency and LD dependent genetic architecture. Instead of sampling per-allele SNP effect sizes from a multivariate normal distribution with homogeneous variance across the genome, we assumed that the variance of SNP *j* in population *k* is proportional to 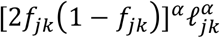, where *f*_*jk*_ and ℓ_*jk*_ are the MAF and LD score of SNP *j* in population *k*, respectively. When *α* < 0, variants with lower MAF and variants located in lower LD regions tend to have larger effects on the trait^38-40^. We used *α* = −0.25 in this set of simulations, which has been empirically estimated to reflect the relationship between effect size and allele frequency^38^. This *α* value produced approximately a 4-fold difference in the variance of per-allele effect size for both high-frequency vs. low-frequency variants and high-LD vs. low-LD variants included in the simulations; (ix) varying hyper-parameters in the continuous shrinkage prior (*a* = 0.5, *b* = 0.5 vs. *a* = 1.0, *b* = 0.5 vs. *a* = 1.5, *b* = 0.5 vs. *a* = 1.0, *b* = 1.0).

### UKBB, BBJ, PAGE and TWB analysis

<u>Discovery data</u>: We downloaded GWAS summary statistics from UKBB^28^, BBJ^29^ and PAGE^31^ (Data availability). We selected 33 quantitative traits that were available in both UKBB and BBJ, among which 14 were also available in PAGE (Supplementary Table 10). We used 1KG EUR and EAS samples as the LD reference panel for UKBB and BBJ summary statistics, respectively, when constructing PRS. The PAGE study was largely comprised of African American and Hispanic/Latino samples, for which we used the 1KG AMR reference panel as an approximation in the PRS analyses. <u>UKBB target data</u>: All UKBB target samples are unrelated UKBB individuals that are non-overlapping and unrelated with the UKBB GWAS sample. To perform population assignment on the UKBB samples, we selected variants that are available in both 1KG and the UKBB genotyped dataset, and removed variants meeting one of the following criteria in 1KG: (i) strand ambiguous; (ii) located on sex chromosomes or in long-range LD regions (chr6: 25-35Mb; chr8: 7-13Mb); (iii) call rate <0.98; and (iv) MAF <0.05. We performed LD pruning on the remaining variants in 1KG using PLINK^49^ (--indep-pairwise 100 50 0.2), yielding 149,501 largely independent, high-quality common variants. We then conducted principal component analysis using these LD-pruned SNPs in 1KG samples, and projected SNP loadings onto UKBB samples with scale appropriately adjusted. Using 1KG as the reference, we trained a random forest model to predict the 5 super-population labels (AFR, AMR, EAS, EUR, SAS) using the top 6 PCs, and applied the trained random forest classifier to UKBB samples to predict the genetic ancestry of each UKBB participant. We retained UKBB samples that can be assigned to one of the super-populations with a predicted probability >90%. For each population in UKBB, we selected a set of unrelated individuals and performed sample-level quality control (QC) by removing individuals meeting one of the following criteria: (i) mismatch between self-reported and genetically inferred sex; (ii) missingness or heterozygosity outliers; and (iii) sex chromosome aneuploidy. For the validation and testing of PRS in the EUR population, we used non-British EUR samples that are unrelated to the White British samples included in Neale Lab UKBB GWAS. Lastly, we converted imputed dosage data into hard coded genotypes using PLINK 2.0 with default parameters (i.e., dosage was rounded to the nearest hardcall when the distance was no great than 0.1; otherwise a missing hardcall was saved), and performed variant-level QC within each target population by removing variants meeting one of the following criteria: (i) call rate <0.98; (ii) MAF <0.01; (iii) Hardy-Weinberg equilibrium test *p*-value <10^−10^; and (iv) imputation INFO score <0.8. The final target dataset included 7,507 AFR, 687 AMR, 2,181 EAS, 14,085 EUR and 8,412 SAS individuals, with 12,886,200, 8,593,932, 6,506,126, 8,211,053 and 8,032,121 variants, respectively. <u>TWB target data</u>: The Taiwan Biobank (TWB)^32^ is a prospective cohort study of the Taiwanese population. Participants were 30 to 70 years old at recruitment. Among the 33 quantitative traits examined in UKBB, we identified 21 traits that were also available in TWB. We used 14,232 samples genotyped on the TWBv2 custom array, the same dataset used in the PRS analysis of our recent TWB quantitative trait GWAS study, to evaluate the predictive performance of different polygenic prediction methods. Following the same sample-level and variant-level QC procedures used in the UKBB analysis, the final analytic sample included 10,149 unrelated individuals of EAS ancestry that had complete data across the 21 traits. Detailed information on the sample characteristics, collection of phenotypes, and the QC and imputation of the genotype data can be found in Chen et al.^32^

### Heritability and cross-population genetic correlation

Heritability of each trait in UKBB, BBJ and PAGE was estimated using LD score regression^50^ with ancestry-matched LD reference panels. We calculated the genetic correlation between UKBB, BBJ and PAGE using POPCORN^17^ with default parameters. POPCORN requires the LD score^50^ and cross-covariance score as the input. We used the pre-computed EUR-EAS scores (available from the POPCORN website), and computed EUR-AFR and EAS-AFR scores using the ‘compute’ function provided by POPOCORN on 1KG Phase 3 samples.

### Schizophrenia datasets

Schizophrenia data used in this study is summarized in Supplementary Table 14. PGC wave 2 schizophrenia GWAS summary statistics^33^ were used as the European discovery dataset. Except for one cohort (TMIM1), EAS samples used as discovery and target datasets were described in Lam et al.^16^ TMIM1 was recruited from multiple university hospitals and local hospitals in Japan. Patients were diagnosed according to the Diagnostic and Statistical Manual of Mental Disorders, 4th Edition (DSM-IV) with consensus from at least two experienced psychiatrists. All patients agreed to participate in the study and provided written informed consent. The study was approved by the Institutional Review Boards of the Tokyo Metropolitan Institute of Medical Science and all affiliated institutions. DNA samples were genotyped on the Illumina Infinium Global Screening Array-24 v1.0 (GSA) BeadChip at the Broad Institute, using standard reagents and HTS workflow procedures. GWAS QC and imputation were performed using Ricopili^51^ with default parameters. When used as a target cohort, SNPs were further filtered by imputation INFO score <0.9 and MAF <0.01.

## SUPPLEMENTARY TABLES

**Supplementary Table 1:** Prediction accuracy of different polygenic prediction methods in the primary simulation setting. Phenotypes were simulated using 1% of randomly sampled causal variants (shared across populations), a cross-population genetic correlation of 0.7, and SNP heritability of 50%. PRS were trained using 100K EUR samples and 20K non-EUR (EAS or AFR) samples.

**Supplementary Table 2:** Prediction accuracy of different polygenic prediction methods across different genetic architectures. Phenotypes were simulated using 0.1%, 1% or 10% of randomly sampled causal variants (shared across populations), a cross-population genetic correlation of 0.7, and SNP heritability of 50%. PRS were trained using 100K EUR samples and 20K non-EUR (EAS or AFR) samples.

**Supplementary Table 3:** Prediction accuracy of different polygenic prediction methods across different cross-population genetic correlations. Phenotypes were simulated using 1% of randomly sampled causal variants (shared across populations), a cross-population genetic correlation of 0.4, 0.7 or 1.0, and SNP heritability of 50%. PRS were trained using 100K EUR samples and 20K non-EUR (EAS or AFR) samples.

**Supplementary Table 4:** Prediction accuracy of different polygenic prediction methods across different discovery GWAS sample sizes. Phenotypes were simulated using 1% of randomly sampled causal variants (shared across populations), a cross-population genetic correlation of 0.7, and SNP heritability of 50%. PRS were trained using 50K EUR and 10K non-EUR (EAS or AFR) samples, 100K EUR and 20K non-EUR samples, 200K EUR and 40K non-EUR samples, or 300K EUR and 60K non-EUR samples.

**Supplementary Table 5:** Prediction accuracy of different polygenic prediction methods across different ratios of EUR vs. non-EUR GWAS sample sizes. Phenotypes were simulated using 1% of randomly sampled causal variants (shared across populations), a cross-population genetic correlation of 0.7, and SNP heritability of 50%. PRS were trained using 120K EUR samples without non-EUR samples, 100K EUR and 20K non-EUR (EAS or AFR) samples, 80K EUR and 40K non-EUR samples, or 60K EUR and 60K non-EUR samples.

**Supplementary Table 6:** Prediction accuracy of different polygenic prediction methods across different SNP heritability. Phenotypes were simulated using 1% of randomly sampled causal variants (shared across populations) and a cross-population genetic correlation of 0.7. SNP heritability was fixed at 50% in each population, 50% in the EUR population and 25% in the non-EUR population, or 25% in the EUR population and 50% in the non-EUR population. PRS were trained using 100K EUR samples and 20K non-EUR (EAS or AFR) samples.

**Supplementary Table 7:** Prediction accuracy of different polygenic prediction methods across different proportions of shared causal variants between populations. Phenotypes were simulated using 1% of randomly sampled causal variants. 100%, 70% or 40% of the causal variants were shared across populations. Shared causal variants had a cross-population genetic correlation of 0.7. SNP heritability was fixed at 50%. PRS were trained using 100K EUR samples and 20K non-EUR (EAS or AFR) samples.

**Supplementary Table 8:** Prediction accuracy of different polygenic prediction methods when SNP effect sizes are minor allele frequency (MAF) and LD dependent. Phenotypes were simulated using 1% of randomly sampled causal variants (shared across populations), a cross-population genetic correlation of 0.7, and SNP heritability of 50%. SNP effect sizes were dependent on MAF and LD scores such that SNPs with lower MAF and located in lower LD regions tended to have larger effect sizes. PRS were trained using 100K EUR samples and 20K non-EUR (EAS or AFR) samples.

**Supplementary Table 9:** Prediction accuracy of PRS-CSx across different combinations of hyper-parameters in the continuous shrinkage prior. Phenotypes were simulated using 1% of randomly sampled causal variants (shared across populations), a cross-population genetic correlation of 0.7, and SNP heritability of 50%. Hyper-parameters were set at *a*=1/2, *b*=1/2; *a*=1, *b*=1/2; *a*=3/2, *b*=1/2 or *a*=1, *b*=1. PRS were trained using 100K EUR samples and 20K non-EUR (EAS or AFR) samples.

**Supplementary Table 10:** Quantitative traits examined in this study with GWAS summary statistics from UKBB, BBJ and PAGE, and their heritability and cross-ancestry genetic correlation estimates.

**Supplementary Table 11:** Prediction accuracy of different polygenic prediction methods, trained using the UKBB and BBJ GWAS summary statistics, in different UKBB target populations across 33 quantitative traits.

**Supplementary Table 12:** Prediction accuracy of different polygenic prediction methods, trained using the UKBB and BBJ GWAS summary statistics, in the Taiwan Biobank across 21 quantitative traits.

**Supplementary Table 13:** Prediction accuracy of different polygenic prediction methods, trained using the UKBB, BBJ and PAGE GWAS summary statistics, in different UKBB target populations across 14 quantitative traits.

**Supplementary Table 14:** Information for the schizophrenia case-control cohorts used in this study.

**Supplementary Table 15:** Prediction accuracy of different polygenic prediction methods for schizophrenia risk across 6 East Asian cohorts.

**Supplementary Table 16:** Prevalence of schizophrenia cases at the bottom 2%, 5%, 10% and top 2%, 5%, 10% of the PRS distribution, constructed by different polygenic prediction methods, across 6 EAS cohorts.

## Notes

### Author Declarations

Collection of the UK Biobank (UKBB) data was approved by the UKBB's Research Ethics Committee. UKBB GWAS summary statistics were released by the Neale Lab and are publicly available (http://www.nealelab.is/uk-biobank). UKBB individual-level data used in the present work were obtained under application #32568. Biobank Japan (BBJ) GWAS summary statistics are publicly available on the BBJ website (https://pheweb.jp). The PAGE study GWAS summary statistics are publicly available on the GWAS Catalog (https://www.ebi.ac.uk/gwas/downloads/summary-statistics). Collection of the Taiwan Biobank (TWB) data was approved by the Ethics and Governance Council (EGC) of TWB and the Department of Health and Welfare, Taiwan (Wei-Shu-I-Tzu NO.1010267471). TWB obtained informed consent from all participants for research use of collected data. The access to and the use of TWB data in the present work was approved by the EGC of TWB (approval number: TWBR10907-05) and the Institutional Review Board (IRB) of National Health Research Institutes, Taiwan (approval number: EC1090402-E). Schizophrenia GWAS summary statistics for the European and East Asian populations are publicly available on the Psychiatric Genomics Consortium (PGC) website (https://www.med.unc.edu/pgc/download-results/). The use of schizophrenia individual-level data of East Asian ancestry in the present work was approved by the Stanley Global Asia Initiatives. The following institutions provided ethics oversight for the collection of schizophrenia East Asian data: Domain Specific Review Board (Singapore); University of Hong Kong; Fujita Health University; RIKEN Center for Integrative Medical Sciences; Nagoya University; Osaka University; Niigata University; Bio-X Institutes of Shanghai Jiao Tong University; University Medical Center Utrecht; The University of Western Australia; The University of Indonesia; Peking University Sixth Hospital; National Taiwan University; Samsung Medical Center; Chonnam National University Hospital; Tokyo Metropolitan Institute of Medical Science and affiliated institutions; Harvard Harvard T.H. Chan School of Public Health.

