## Supplementary Information for "Improving Polygenic Prediction in Ancestrally Diverse Populations"

### SUPPLEMENTARY METHODS

PRS-CSx employs the following Bayesian high-dimensional regression framework for $K$ populations:

$$\boldsymbol{y}_k = \boldsymbol{X}_k\boldsymbol{\beta}_k + \boldsymbol{\epsilon}_k, \quad \boldsymbol{\epsilon}_k \sim \mathrm{MVN}\left(\boldsymbol{0}, \sigma_k^2 \boldsymbol{I}\right), \quad p\left(\sigma_k^2\right) \propto \sigma_k^{-2}, \quad k = 1, 2, \cdots, K,$$

where, for each population $k$, $\boldsymbol{y}_k$ is a vector of standardized phenotypes (zero mean and unit variance) from $N_k$ individuals, $\boldsymbol{X}_k$ is an $N_k \times M$ matrix of standardized genotypes (each column has zero mean and unit variance), $\boldsymbol{\beta}_k$ is a vector of SNP effect sizes, $\boldsymbol{\epsilon}_k$ is a vector of normally distributed non-genetic effects with variance $\sigma_k^2$, and $\boldsymbol{I}$ is an identify matrix. For SNP $j$ in population $k$, a continuous shrinkage prior is placed on its effect size $\beta_{jk}$, which can be represented as global-local scale mixtures of normals:

$$\beta_{jk} \sim \mathrm{N}\left(0, \frac{\sigma_k^2}{N_k}\psi_j\right), \quad \psi_j \sim \mathrm{G}\left(a, \delta_j\right), \quad \delta_j \sim \mathrm{G}(b, \phi),$$

where $\phi$ is a global shrinkage parameter shared across all SNPs that models the overall sparseness of the genetic architecture, and $\psi_j$ is a local, SNP-specific shrinkage parameter that is adaptive to marginal GWAS associations. Note that both $\phi$ and $\psi_j$ do not depend on $k$, and thus the continuous shrinkage prior is shared across populations.

The full conditional distributions for unknown model parameters are analytically tractable. Let $\mathrm{MVN}(\boldsymbol{\mu}, \boldsymbol{\Sigma})$ denote the multivariate normal distribution with mean $\boldsymbol{\mu}$ and covariance matrix $\boldsymbol{\Sigma}$; $\mathrm{G}(\zeta, \eta)$ and $\mathrm{iG}(\zeta, \eta)$ denote the gamma distribution and inverse gamma distribution, respectively, with probability density functions

$$f_{\mathrm{G}}(x; \zeta, \eta) = \frac{\eta^\zeta}{\Gamma(\zeta)} x^{\zeta-1} \exp(-\eta x), \quad f_{\mathrm{iG}}(x; \zeta, \eta) = \frac{\eta^\zeta}{\Gamma(\zeta)} x^{-\zeta-1} \exp\left(-\frac{\eta}{x}\right), \quad x > 0, \quad \zeta > 0, \quad \eta > 0,$$

where $\Gamma(\cdot)$ is the gamma function; and $\mathrm{giG}(\lambda, \rho, \chi)$ denote the three-parameter generalized inverse Gaussian distribution with density function

$$f_{\mathrm{giG}}(x; \lambda, \rho, \chi) = \frac{(\rho/\chi)^{\lambda/2}}{2K_\lambda\left(\sqrt{\rho\chi}\right)} x^{\lambda-1} \exp\left\{-\frac{1}{2}\left(\rho x + \frac{\chi}{x}\right)\right\}, \quad x > 0, \quad \rho > 0, \quad \chi > 0,$$

where $K_\lambda$ is the modified Bessel function of the second kind. In addition, let $\widehat{\boldsymbol{\beta}}_k = \boldsymbol{X}_k^T\boldsymbol{y}_k/N_k$ denote the marginal least squares effect size estimates from the GWAS summary statistics for population $k$, $\boldsymbol{D}_k = \boldsymbol{X}_k^T\boldsymbol{X}_k/N_k$ denote the LD matrix for population $k$, $\boldsymbol{\Psi} = \mathrm{diag}\{\psi_1, \psi_2, \cdots, \psi_M\}$, where $M$ is the total number of unique SNPs across populations, and $k_j$ denote the number of populations in which SNP $j$ is present. The Gibbs sampler for the PRS-CSx model involves the following steps in each Markov Chain Monte Carlo (MCMC) iteration:

- Update $\boldsymbol{\beta}_k$:

$$[\,\boldsymbol{\beta}_k \mid \sigma_k^2, \boldsymbol{\Psi}, \widehat{\boldsymbol{\beta}}_k, \boldsymbol{D}_k\,] \sim \mathrm{MVN}(\boldsymbol{\mu}_k, \boldsymbol{\Sigma}_k), \quad \boldsymbol{\mu}_k = \frac{N_k}{\sigma_k^2}\boldsymbol{\Sigma}_k\widehat{\boldsymbol{\beta}}_k, \quad \boldsymbol{\Sigma}_k = \frac{\sigma_k^2}{N_k}\left(\boldsymbol{D}_k + \boldsymbol{\Psi}^{-1}\right)^{-1},$$

- Update $\sigma_k^2$:

$$[\,\sigma_k^2 \mid \boldsymbol{\beta}_k, \boldsymbol{\Psi}, \widehat{\boldsymbol{\beta}}_k, \boldsymbol{D}_k\,] \sim \mathrm{iG}\left(\frac{N_k + M}{2}, \frac{N_k}{2}\left[1 - 2\boldsymbol{\beta}_k^T\widehat{\boldsymbol{\beta}}_k + \boldsymbol{\beta}_k^T\left(\boldsymbol{D}_k + \boldsymbol{\Psi}^{-1}\right)\boldsymbol{\beta}_k\right]\right),$$

- Update $\psi_j$:

$$[\ \psi_j \mid \beta_{jk}, \sigma_k^2, \delta_j\ ] \sim \mathrm{giG}\left(a - \frac{k_j}{2}, 2\delta_j, \sum\nolimits_k \frac{N_k}{\sigma_k^2} \beta_{jk}^2\right),$$

- Update $\delta_j$:

$$[\ \delta_j \mid \psi_j\ ] \sim \mathrm{G}\big(a + b, \psi_j + \phi\big).$$

### SUPPLEMENTARY FIGURES

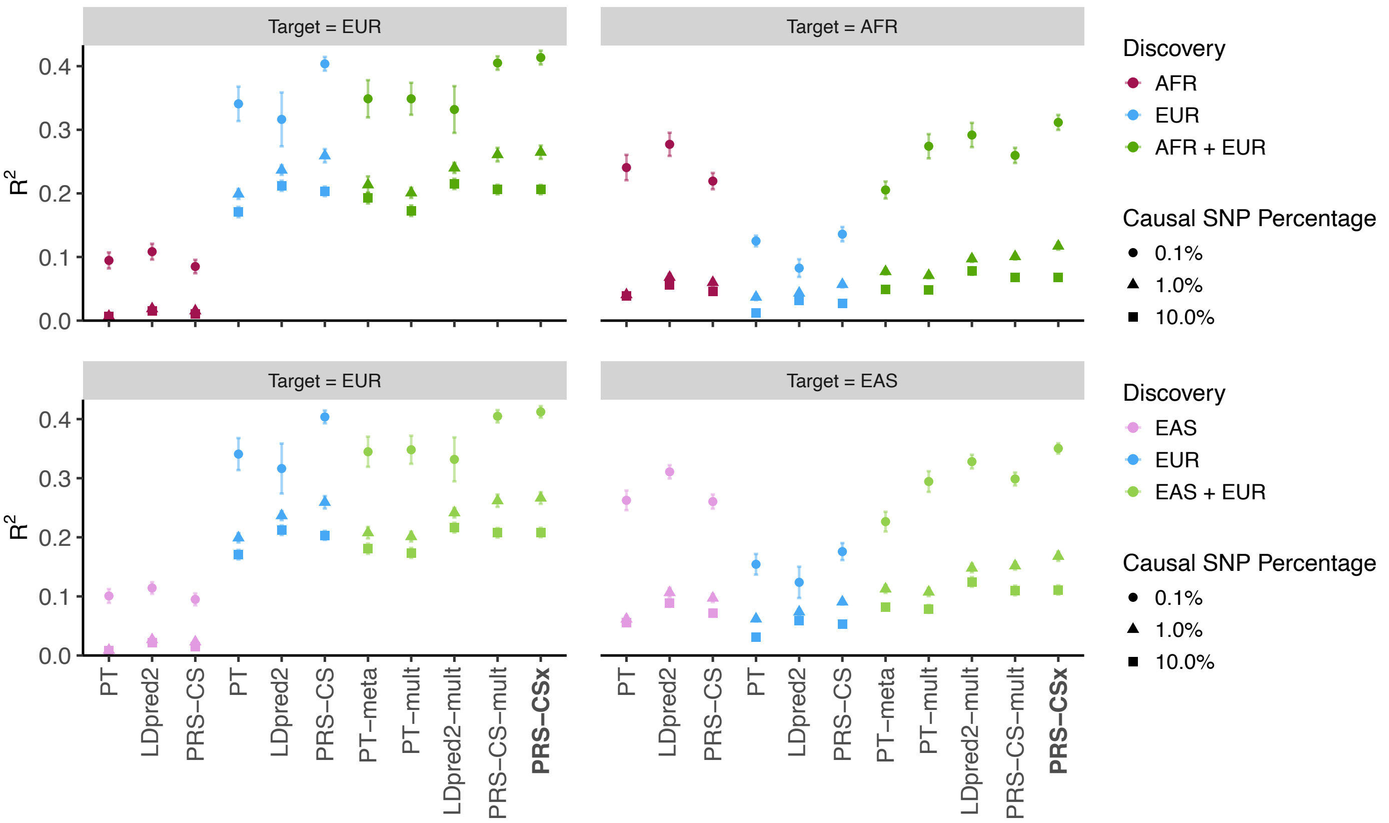


**Supplementary Figure 1:** Prediction accuracy of different polygenic prediction methods across different genetic architectures. Phenotypes were simulated using 0.1%, 1% or 10% of randomly sampled causal variants (shared across populations), a cross-population genetic correlation of 0.7, and SNP heritability of 50%. PRS were trained using 100K EUR samples and 20K non-EUR (EAS or AFR) samples. Numerical results are reported in Supplementary Table 2.

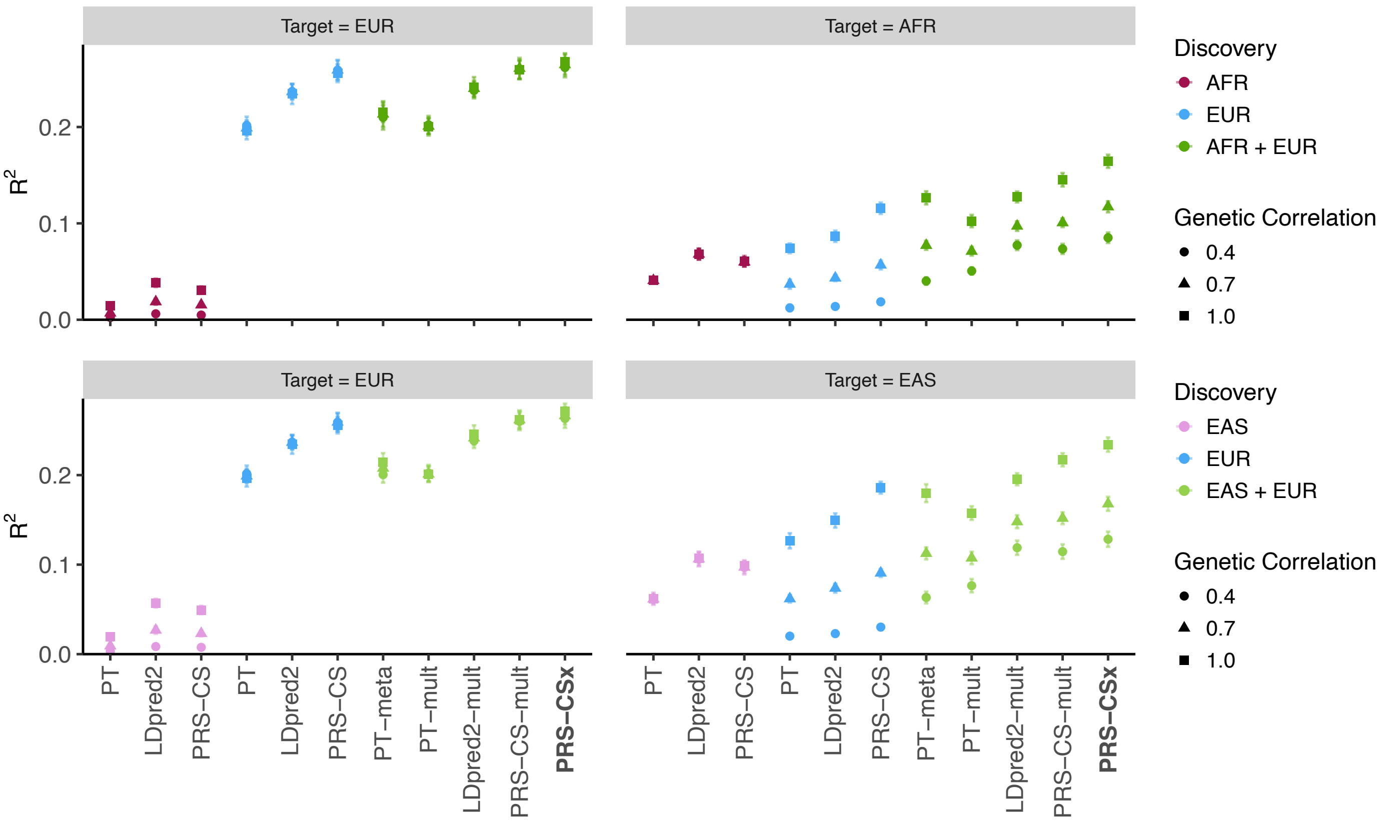


**Supplementary Figure 2:** Prediction accuracy of different polygenic prediction methods across different cross-population genetic correlations. Phenotypes were simulated using 1% of randomly sampled causal variants (shared across populations), a cross-population genetic correlation of 0.4, 0.7 or 1.0, and SNP heritability of 50%. PRS were trained using 100K EUR samples and 20K non-EUR (EAS or AFR) samples. Numerical results are reported in Supplementary Table 3.

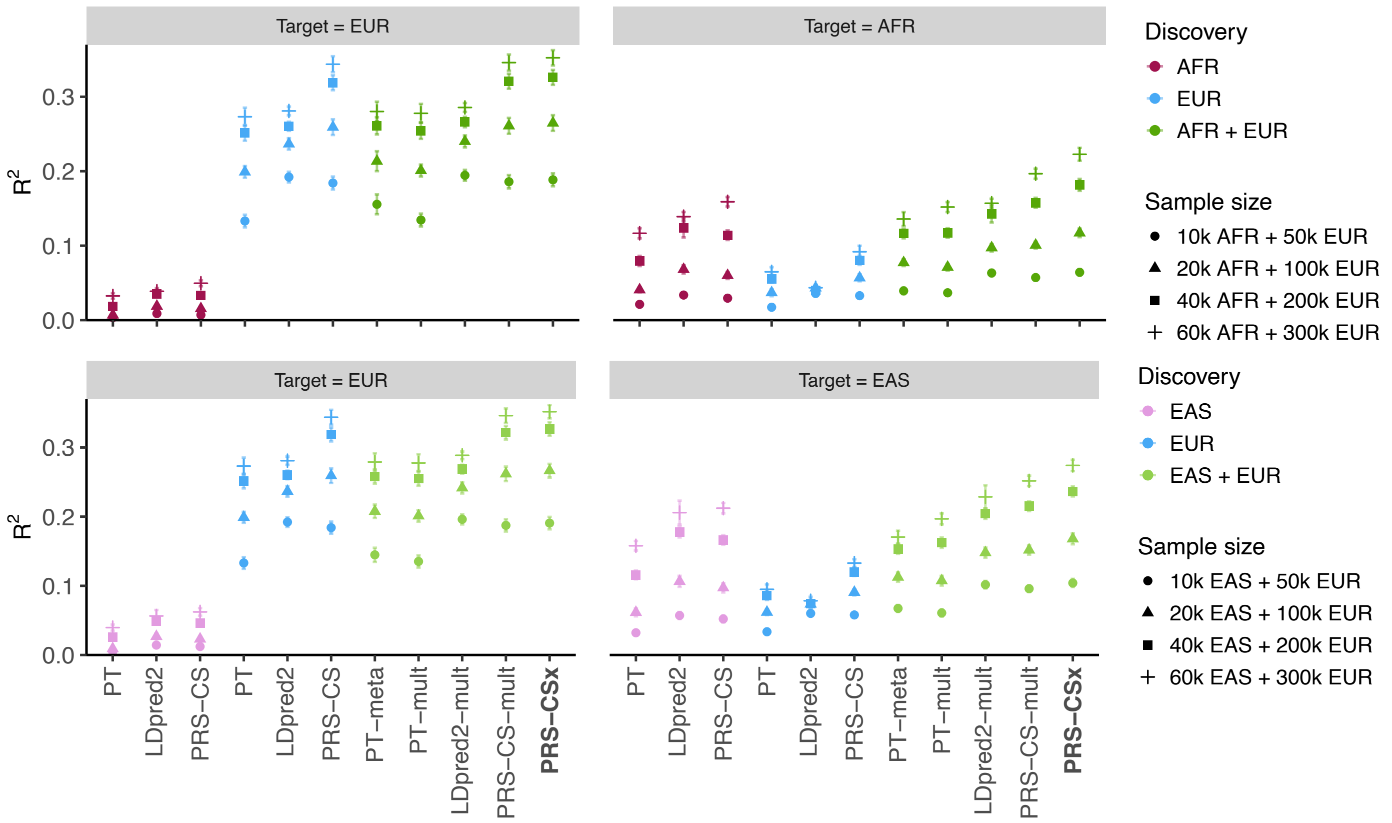


**Supplementary Figure 3:** Prediction accuracy of different polygenic prediction methods across different discovery GWAS sample sizes. Phenotypes were simulated using 1% of randomly sampled causal variants (shared across populations), a cross-population genetic correlation of 0.7, and SNP heritability of 50%. PRS were trained using 50K EUR and 10K non-EUR (EAS or AFR) samples, 100K EUR and 20K non-EUR samples, 200K EUR and 40K non-EUR samples, or 300K EUR and 60K non-EUR samples. Numerical results are reported in Supplementary Table 4.

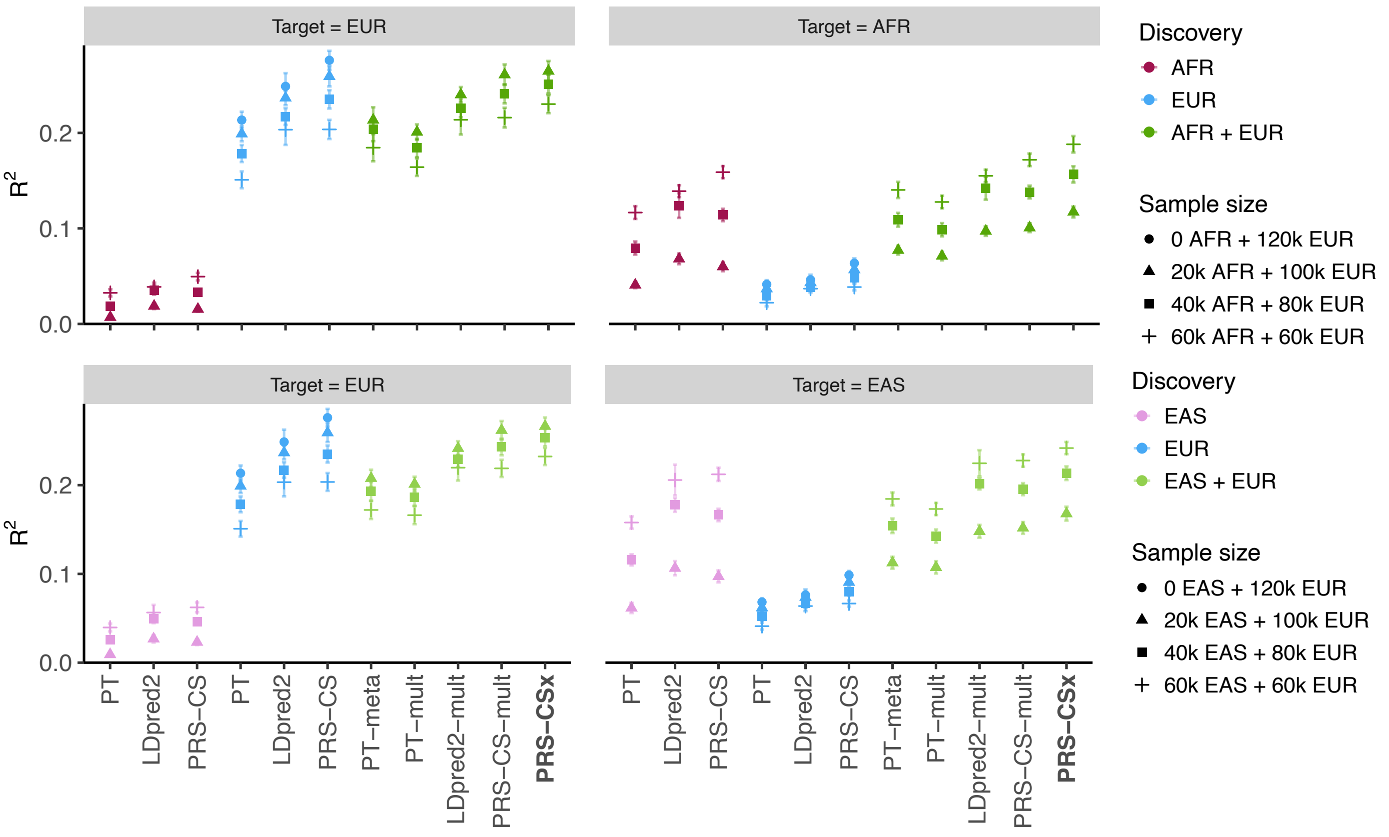


**Supplementary Figure 4:** Prediction accuracy of different polygenic prediction methods across different ratios of EUR vs. non-EUR GWAS sample sizes. Phenotypes were simulated using 1% of randomly sampled causal variants (shared across populations), a cross-population genetic correlation of 0.7, and SNP heritability of 50%. PRS were trained using 120K EUR samples without non-EUR samples, 100K EUR and 20K non-EUR (EAS or AFR) samples, 80K EUR and 40K non-EUR samples, or 60K EUR and 60K non-EUR samples. Numerical results are reported in Supplementary Table 5.

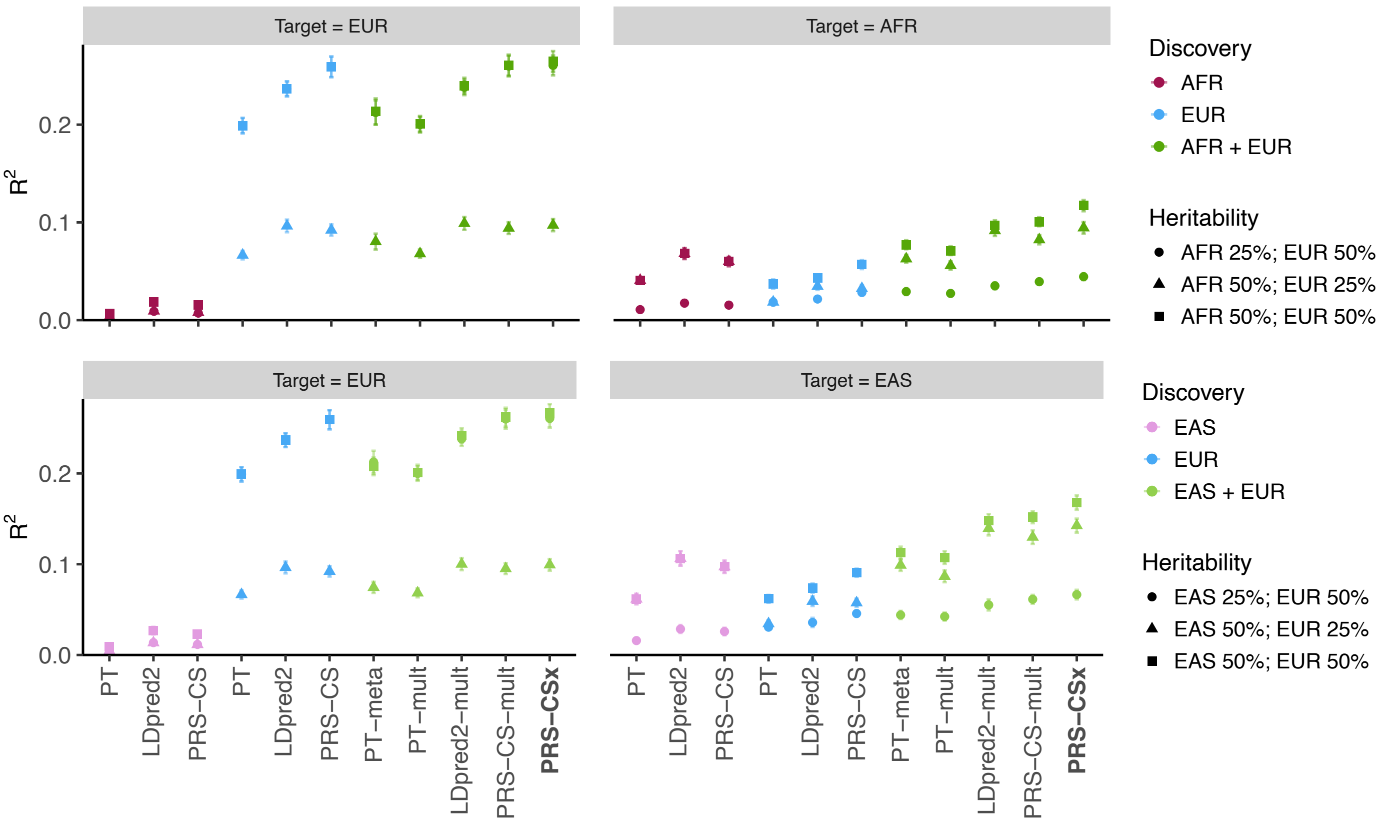


**Supplementary Figure 5:** Prediction accuracy of different polygenic prediction methods across different SNP heritability. Phenotypes were simulated using 1% of randomly sampled causal variants (shared across populations) and a cross-population genetic correlation of 0.7. SNP heritability was fixed at 50% in each population, 50% in the EUR population and 25% in the non-EUR population, or 25% in the EUR population and 50% in the non-EUR population. PRS were trained using 100K EUR samples and 20K non-EUR (EAS or AFR) samples. Numerical results are reported in Supplementary Table 6.

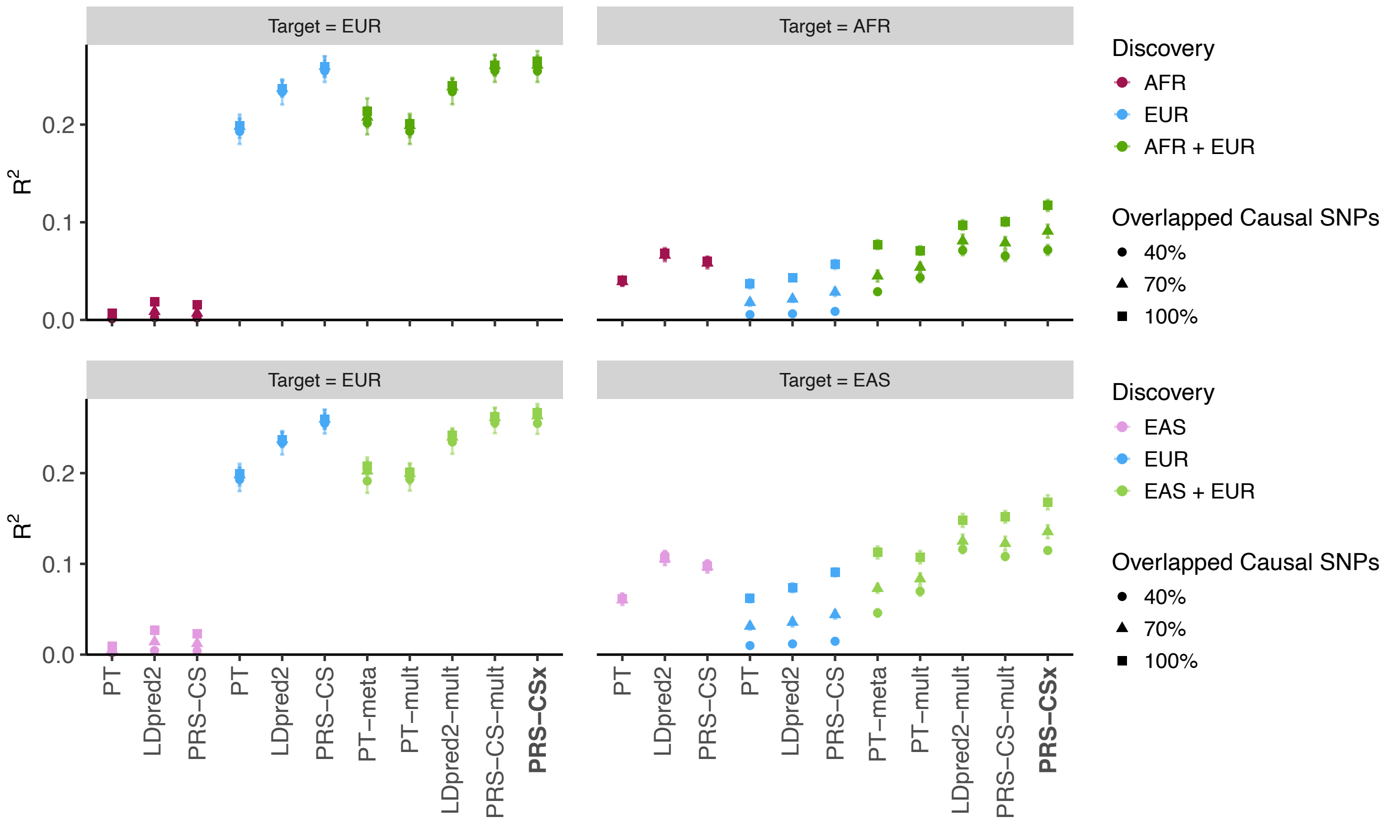


**Supplementary Figure 6:** Prediction accuracy of different polygenic prediction methods across different proportions of shared causal variants between populations. Phenotypes were simulated using 1% of randomly sampled causal variants. 100%, 70% or 40% of the causal variants were shared across populations. Shared causal variants had a cross-population genetic correlation of 0.7. SNP heritability was fixed at 50%. PRS were trained using 100K EUR samples and 20K non-EUR (EAS or AFR) samples. Numerical results are reported in Supplementary Table 7.

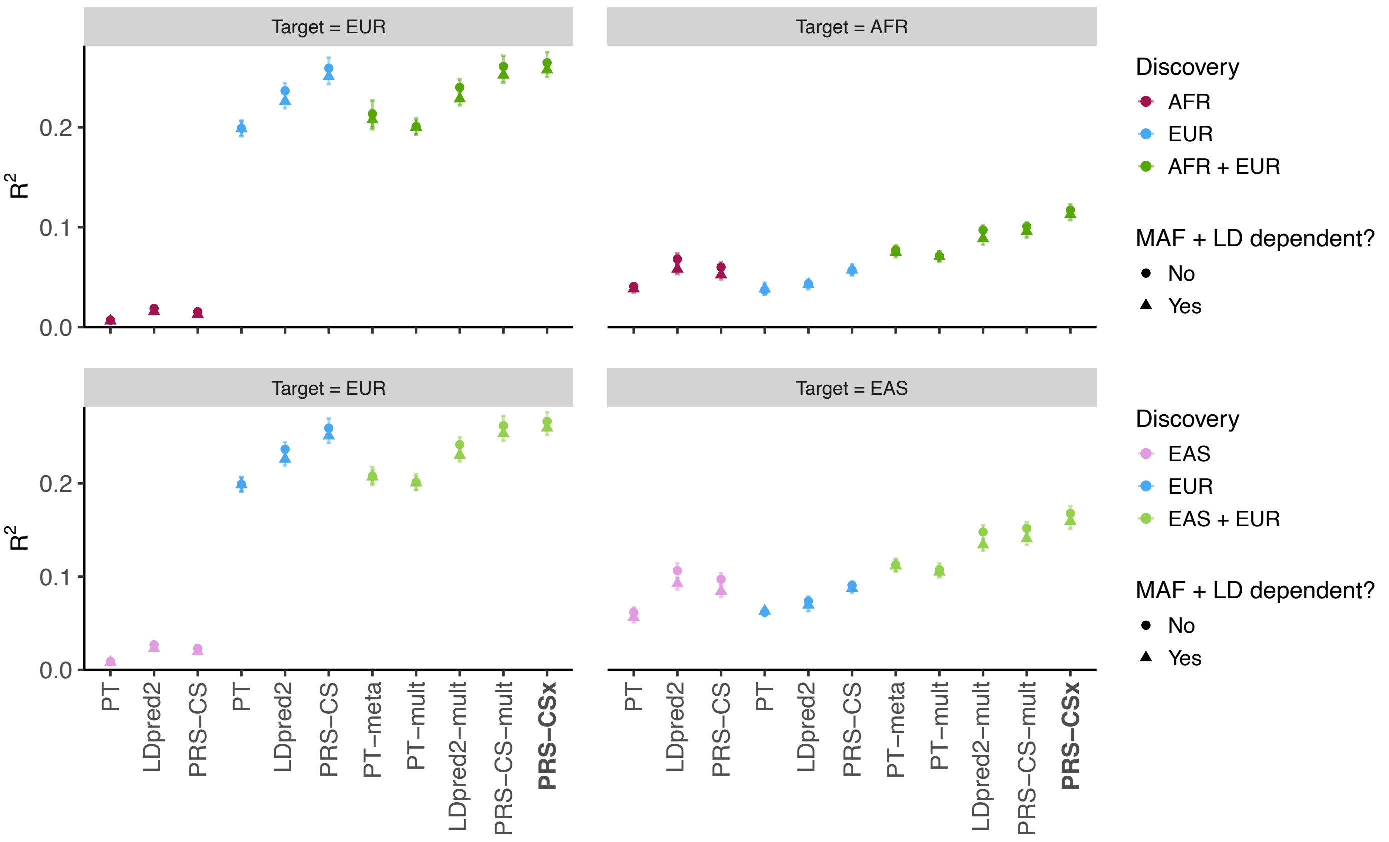


**Supplementary Figure 7:** Prediction accuracy of different polygenic prediction methods when SNP effect sizes are minor allele frequency (MAF) and LD dependent. Phenotypes were simulated using 1% of randomly sampled causal variants (shared across populations), a cross-population genetic correlation of 0.7, and SNP heritability of 50%. SNP effect sizes were dependent on MAF and LD scores such that SNPs with lower MAF and located in lower LD regions tended to have larger effect sizes. PRS were trained using 100K EUR samples and 20K non-EUR (EAS or AFR) samples. Numerical results are reported in Supplementary Table 8.

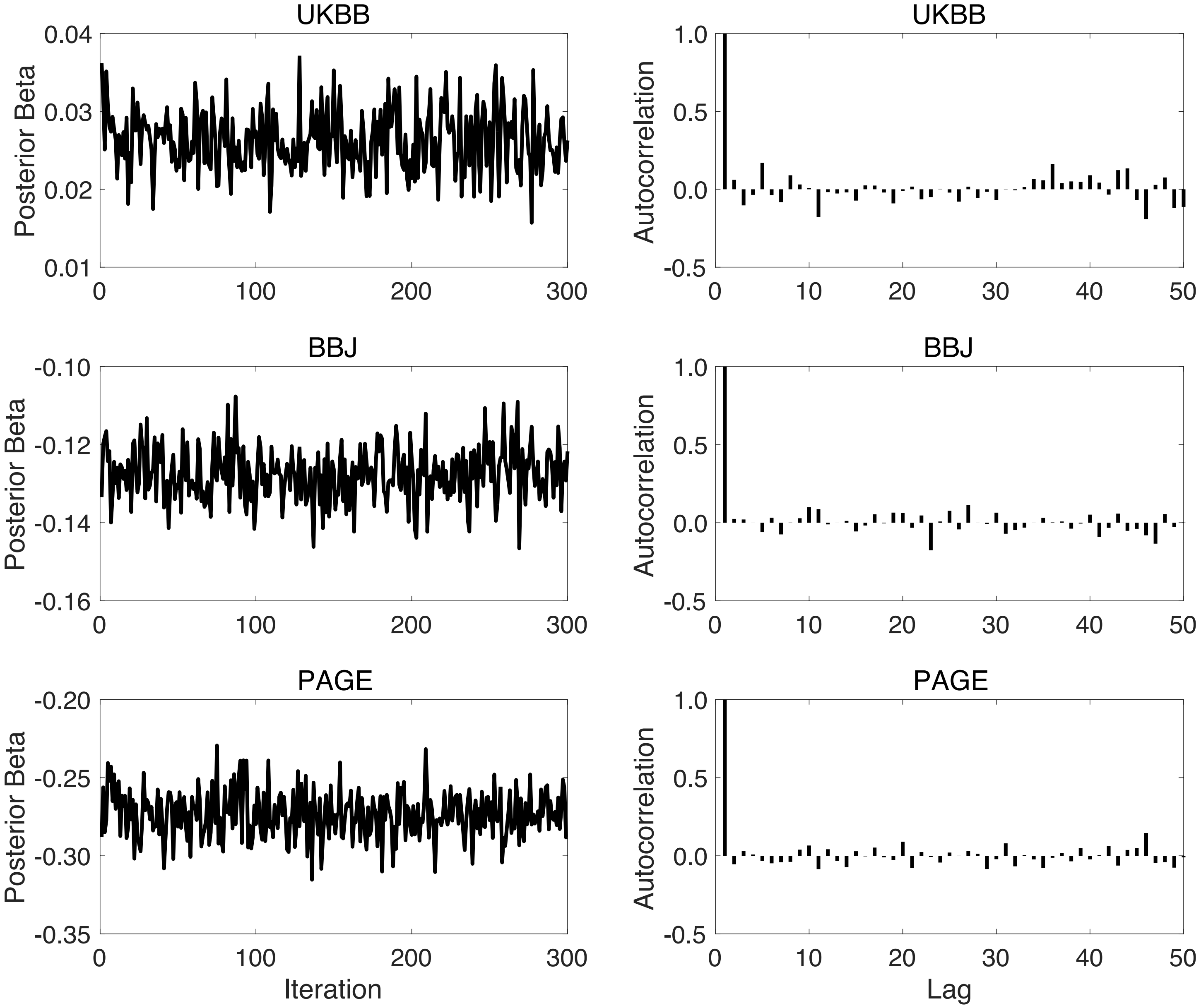


**Supplementary Figure 8:** Trace plots and autocorrelation functions (ACFs) for assessing the convergence and mixing of the Gibbs sampler used in PRS-CSx. Left panels: Trace plots, after discarding the burn-in iterations and thinning the Markov chain by a factor of 5, for the posterior effects of rs7412 on low-density lipoprotein cholesterol when integrating UKBB, BBJ and PAGE GWAS summary statistics using PRS-CSx. Right panels: The autocorrelation functions (ACFs) for the traces shown on the left.
